# Protocol for the Virtual, Nurse-led Early paLlIative Care IntervenTion (ELICIT) Randomized Controlled Trial

**DOI:** 10.1101/2025.04.05.25324800

**Authors:** Vyjeyanthi. S Periyakoil, Charles Von Gunten, Helena Kraemer

## Abstract

**Background:** Though dementia is a serious illness that progresses over many years, little is known about the primary palliative care needs of chronically-ill, community dwelling older adults at risk of already diagnosed with cognitive impairment.

**Objective:** To test the impact of a virtual, nurse-led primary palliative care intervention on older adults at risk of already diagnosed with cognitive impairment.

**Methods:** This randomized clinical trial (RCT) was designed to compare the impact of {usual care} to {usual care plus a virtual, nurse-led, primary palliative care intervention} on community-dwelling, chronically ill older adults with varying degrees of cognitive impairment.

**Results:** We hypothesize that patients in the virtual nurse intervention arm will have their primary palliative care needs identified and fulfilled compared to those in the usual care arm.

**Conclusions:** Identification of the primary palliative care needs of chronically-ill, community dwelling older adults at risk of and/or with MCI, mild or moderate dementia will help refine intervention to effectively provide primary palliative care in a timely manner to these patients.

**Trial Registration:** ClinicalTrials.gov Identifier: NCT03881579

## Introduction

The World Health Organization has declared dementia a public health crisis and issued a call for global nations to make dementia an international public health priority[1]. Though dementia is a serious illness that progresses over many years, little is known about the everyday suffering endured over time by patients in the early stages of the disease as most research focuses on patient who are already institutionalized and at the end of life.

However, patients early in the dementia trajectory are beset with distressing symptoms. An estimated 85%[2]of Persons With Dementia (PWD) live in the community (e.g., private residences or non-nursing home residential care facilities) with unmet needs[3]. Proactive, patient-centered primary palliative care will likely benefit early-stage patients living in the community. Implementing early palliative care will facilitate ongoing assessment and management of the distressing symptoms experienced by PWD from very early in the illness trajectory and result in identification and management of primary palliative care needs and facilitate timely advance care planning[4].

### Preliminary Studies

**Preliminary study 1:** Validating a multi-lingual, low-literacy What Matters Most Letter Advance Directive (WMM): Traditional advance directives are filled with medico-legal jargon and do not capture what-matters-most to patients and their values. Working with diverse patients and families we co-designed the What Matters Most Letter Advance Directive (WMM) in a simple letter format written at a fifth grade reading level in multiple languages (English, Chinese, Hindi, Russian, Spanish, Tagalog, Vietnamese, and Urdu). We tested the WMM through a large, randomized control trial (NCT02799537)[5] – a total of 400 participants participated, with 216 randomized to the WMM and 184 to the State of California’s Advance Directives. Overall, participants preferred the WMM to the California State advance directive (Success Rate Difference [SRD] = 0.46, 95th percentile confidence interval [CI] 0.36-0.56, p<0.001). The participants felt that the WMM was easier to read and understand (SRD=0.56 [CI]=0.47-0.65), p<0.001); better reflected what matters most to them (SRD =0.39 [CI]: 0.29-0.48, p<0.001,); helped stimulate their thinking about the types of treatments they wanted at the End-of-life (SRD=0.32 [CI]: 0.23-0.42), p<0.001); allowed them to describe how they made medical decisions in their family (SRD=0.31 [CI]:0. 21-0.40), p<0.001); and could help their doctor(s) (SRD=0.24, [CI]: 0.13-0.34, p<0.001), and their families (SRD=0.19, [CI]: 0.08-0.28), p<0.001) understand their treatment preferences. 33.5% of the RCT participants were Hispanic/Latino Americans and they preferred to complete the WMM over the California State advance directive as it was written in simple language.
**Preliminary study 2:** We have demonstrated that the *<u>WMM facilitates patient-proxy</u> <u>collaboration in completing advance directives.</u>* We recruited 81 seriously ill patients[6] admitted to the Stanford Hospital between 3/1/2016 to 2/28/2017 and implemented the following study: The patient (L1) and proxy (L2) first completed the WMM individually. Next, they discussed the contents of the WMM and collaborated to complete, sign and file an advance directive. <u>The WMM allows</u> <u>patients to choose from a menu of various common end of life treatment</u> <u>options. Quantitative data analyses revealed that initially, patients and their proxies had</u> <u>varying degrees</u> <u>of agreement on the specific treatment choices:</u> 79% agreement on the question about cardiac resuscitation attempts, 73% agreement about dialysis, 65% agreement for artificial nutrition and hydration, 56% agreement on the question about ventilatory support. 84% agreement about wanting to be pain-free, 76% agreement about wanting to be sedated if their pain and symptoms became difficult to control (76%). 63% agree about not wanting to spend their last days in the hospital, 64% agreement about wanting to die gently and naturally and 54% agreement about wanting hospice care. However, when they discussed their differences, the patient and proxy were able to align closely and complete a consensus WMM, which was almost always in favor of the patient’s decisions. All patients and their proxies reported that they very much preferred the WMM to traditional advance directives, so much so that Stanford Hospital has implemented the WMM as standard practice.
**Preliminary study 3:** Providers prefer the WMM over traditional advance directives[7]: We performed a prospective, cluster-randomized, controlled trial with five general medicine ward teams at Stanford Hospital in 2016. Patients admitted to two control teams received usual care, including the standard protocol for offering the California State advance directive. In addition to usual care, patients admitted to three intervention teams were offered the WMM. Doctors in each of the teams were surveyed regarding their self-assessed understanding of their patients’ goals of care as well as the usefulness of the letter. Chi square and T tests were used to compare the two groups. Doctors on the control teams completed 93% of the surveys and those on the intervention teams completed 100% of the surveys. On a five-point Likert scale, intervention team providers rated themselves more highly than the control providers with regards to (i) understanding of “what matters most” to their patients (mean score 3.96 versus 2.47 respectively, p<0.05) and (ii) their feeling of preparedness to guide their patient’s proxy (mean score 3.63 versus 2.67 respectively, p<0.05). Compared to the California State advance directive, the providers found the WMM most helpful 92% of the time, reporting that it (i) improved understanding of goals of care; (ii) would be useful in guiding end-of-life discussions; (iii) captured end-of-life preferences in the patient’s own words; and (iv) helped clarify patient values and family dynamics. Intervention team providers were also more likely than usual care providers to report knowing their patients’ preferred site of death (79% versus 20% of responses respectively, p<0.05). Thus, the WMM is very helpful to the doctors who are better able to understand their patients’ life goals and values and improve shared decision-making around goals of care and advance care planning. This study won a research poster award from the American College of Physicians (2016) and was chosen as one of the best oral abstract presentations by the Society of Hospital Medicine (2017). The WMM won an innovation award (2015) from the American Medical Association and the American College of Physicians created a Steps Forward module on the WMM and provides implementation support. In the proposed study the research nurse will use the WMM (English and Spanish) to help patients and families identify and document what matters most to them.

### Validating the training provided to the ELICIT trial research nurse interventionists

Dr. Patel, a collaborator of Dr. Periyakoil led an RCT “Engagement of Patients with Advanced Cancer Trial (NCT02966509)”[8] where the project coordinators (lay health workers) were trained by Dr. Periyakoil’s lab through our iSAGE training program (a hybrid training program with online training provided through the Stanford Canvas Learning Management System followed by immersive training and practice with feedback with standardized patients and receive feedback to refine their communication skills. This study was published in JAMA Oncology (PMID: 30054634 PMCID: PMC6233780[9]). In the proposed ELICIT RCT, the research nurses will be trained using this validated training approach and will receive consultation and supervision about all their patients with Dr. Periyakoil.

### Methods

#### STUDY DESIGN

Recognizing that the key components of primary palliative care include on the four identified primary palliative care domains: (1) assessing and assisting with palliation of distressing physical symptoms; (2) assessing and supporting psychological, social, cultural, and spiritual aspects of care; (3) establishing goals of care, identification of a surrogate decision maker and completion of advance care planning documentation; and (4) providing care coordination support, we have designed, refined, and piloted an intervention to be used in the proposed Randomized Clinical Trial (RCT). We will recruit 200 chronically-ill, community dwelling older adults who have undergone full assessment by a multi-disciplinary neurology team and are at risk for or already have cognitive impairment. They will be randomized 100 each to usual care (UC) or usual care plus an early primary palliative care intervention (EPC) is to be delivered by a nurse over a twelve-month period. The virtual EPC will include one nurse-led palliative consult in an initial encounter (up to 2 hours) and followed by 11 monthly 30-minute monthly encounters plus usual care.

## RESULTS

### Primary Outcome Measures

1. To identify the number of participants who express supportive care needs in both arms. Only the participants randomized to the intervention arm will receive the nurse-led supportive care intervention (one session per month over a twelve-month period).
2. Completion and documentation of advance directives (AD) and the Physicians Orders for Life Sustaining Treatment (POLST) in the electronic health records.

[Time Frame: Day 0, 4 months, one year]

We hypothesize that compared to the control arm, many more patients in the intervention arm will (a) express supportive needs and have them palliated by the study nurses and (b) complete advance directives and POLST documentation. In both arms the investigators will track the completion, signage and documentation of advance directives and the POLST. AD has to be signed by the patient or proxy and witnessed by two qualifying witnesses. The POLST has to be signed by the patient/proxy and the patient’s doctor. Both forms have to be uploaded into the electronic health records.

[Time Frame: Day 0, 4 months, one year]

### Secondary Outcome Measures

1. Changes in Edmonton Symptoms Assessment Scale (ESAS) scores: The ESAS is designed to assist in the assessment of nine symptoms common in patients: pain, tiredness, nausea, depression, anxiety, drowsiness, appetite, wellbeing and shortness of breath, (there is also a line labeled “Other Problem“). The severity at the time of assessment of each symptom is rated from 0 to 10 on a numerical scale, 0 meaning that the symptom is absent and 10 that it is of the worst possible severity. The investigators will assess change in ESAS score of all participants at baseline(Day 0), months 4, 12 and 18. The investigators hypothesize that patients with cognitive impairment will have higher scores than those with normal cognition. The investigators will also determine changes in the ESAS score (if any) over time. [Time Frame: Day 0, 4 months, one year]
2. Changes in Patient Activation Measure (PAM) over time: the PAM is a 3-item survey that assesses a person’s underlying knowledge, skills and confidence integral to managing his or her own health and healthcare. PAM segments individuals into one of four activation levels along an empirically derived 100-point scale. Each level provides insight into an extensive array of health-related characteristics, including attitudes, motivators, and behaviors. Individuals in the lowest activation level do not yet understand the importance of their role in managing their own health, and have significant knowledge gaps and limited self-management skills. Individuals in the highest activation level are proactive with their health, have developed strong self-management skills, and are resilient in times of stress or change. PAM measures patient activation and agency: The investigators will assess changes in PAM scores (if any) over time. [Time Frame: Day 0, 4 months, one year]

**Quality of Life Alzheimer’s Disease (QOL-AD): QOL-AD:** The QoL-AD is a valid and reliable scale which is brief (13 items), has good content validity, good criterion concurrent validity and construct validity. Interrater reliability was strong with all Cohen’s kappa values >0.70. Internal consistency was good with a Cronbach’s alpha of 0.82. Some people with severe dementia and a Mini-Mental State Examination score as low as 3 were able to satisfactorily complete the QoL-AD. QOL-AD scores

[Time Frame: Day 0, 4 months, one year]

## TRAINING OF NURSE INTERVENTIONISTS

Nurses experienced in caring for patients with chronic illness will be recruited. All nurses were trained to use the intervention measures by the PI. Additionally, all nurses will complete a canvas course as described above. All will have access to the training modules. Ongoing training for skill building and education will be provided through weekly team meetings (Tuesday afternoons).

## ELICIT INTERVENTION

| Intervention Session | Type and duration | Content and format |
| --- | --- | --- |
| Session 1 (S1) | Secure initial video encounter | Complete assessment of the patient's primary palliative care needs using the measures shown below and assess patient's preferences for care. Introduce advance care planning and goals of care discussions.<br>Post S1, the nurse presents the patient information and stated preferences to the study palliative care physician and works to craft a personalized care plan based on the patient's needs. The nurse will then circle back to the patient to complete tasks related to the primary palliative care issues identified. |
| Sessions 2-12 | Secure monthly video encounter or audio encounter (based on participant preference) | The nurse will review the patient's situation over the past month, identify any concerns. S/he will complete the distress thermometer and use it to explore any underlying issues to be palliated and discuss those with the study palliative care physician. Any necessary measures will be taken to fulfill the patient's primary palliative care needs throughout the entire period of the intervention (12 months)<br>The nurse will continue the discussion about what matters most to the patient and provide assistance in completing the WMM and the POLST form. If the WMM is signed by the patient, the nurse will help identify two valid adult witnesses to witness the WMM form. The signed WMM form is scanned into the patient's electronic medical records.<br>If the patient is willing, the nurse discusses the POLST form choices with the patient and helps them complete the form. The patient is encouraged to discuss the POLST with their primary care doctor and finalize their choices. Once the patient and the primary care doctor sign the POLST form, the nurse will upload the POLST form into the electronic medical records and send an extra |
|  |  | copy to the patient to post on the front door of their refrigerator.<br>At the end of each encounter, the research nurse will work with the patient to identify any issues that need attention and make plans for follow up in the next few days. The nurse will also identify the date for the encounter for the following month until the last intervention month (S12). |
| Study exit information (given at S12) | Secure video encounter or audio encounter (based on participant preference) | The nurse will review the past year and summarize the issues. S/he will provide the contact information for the patient in event they need further assistance. At the end of the intervention, the nurse will mail another copy of their signed WMM and signed POLST to the patient for their records. |

## SUBJECT INVOLVEMENT & CHARACTERISTICS

### Recruitment and Patient Flow

Recruitment of subjects will be through existing institutional mechanisms as follows: After getting approval from the Institutional Review Board (IRB) for this proposed RCT, the patients who meet eligibility criteria will be identified from our REDCAP database. After getting IRB permission, the study team will contact the patient and caregiver by phone to schedule a screening call to determine eligibility. If eligible and if the patient and caregiver consent to participate in the study, we will schedule an appointment with the study evaluators to complete the baseline study measures during a video visit. The project research nurses will render the EPC intervention over a 12-month period. All the intervention sessions will be audio taped for future qualitative analyses.

## INCLUSION CRITERIA

Patients eligible to participate are those who have undergone an evaluation by a multidisciplinary experts who review the patient’s cognitive and functional performance in six areas: memory, orientation, judgment & problem solving, community affairs, home & hobbies, and personal care. Only patients with a composite “Clinical Dementia Rating” (CDR) score of 0 (no symptoms), 0.5 (very mild), 1.0 (mild), 2.0 (moderate) are eligible to participate in the study as long as they are living in the community.

## EXCLUSION CRITERIA

Patients are excluded from the study if they have severe dementia (CDR=3.0) or if they are institutionalized. Outcome measures will be obtained on all participants at baseline, months 4 and 12 by bilingual (English and Spanish) evaluators.

### Proposed Involvement

After obtaining informed consent, patients will be randomized to either the EPC intervention {Usual care + Early Primary Palliative Care} or to UC {Usual Care}. For patients randomized to the EPC, the study nurse(s) will work with the patient/proxy to schedule the session 1 (S1) of intervention as a secured virtual video encounter. Once the first session is complete, sessions 2-12 will be scheduled monthly video or phone nurse encounters. To the extent possible, the same nurse will be assigned to each patient to ensure continuity. At baseline (at the time of study enrollment) and months 4 and 12 a research associate who is blinded to the treatment condition will collect the secondary outcome measures. Long term outcome follow up will be collected at month 18 and thereafter until death or loss to follow up whichever comes first.

### Study Population & Sample Size

200 enrollees who meet inclusion and exclusion criteria to be consented and randomly assigned to intervention {Usual care + Early Primary Palliative Care} of EPC intervention or to {Usual Care} or UC. Consenting caregivers will also be allowed to participate as observers along with the patient. Our IRB requires that the caregivers sign a separate informed consent to participate in the study. Caregivers who do not sign informed consent will not be able to participate.

### Procedure for handling any requests from patients in the intervention arm during the study period

All patients/caregivers in the intervention arm will be contacted by their assigned primary palliative care research nurse on study entry and once monthly during the 12-month intervention period. The assigned primary palliative care research nurse will assess the patient for their primary palliative care needs (using the standard measures listed below), conduct goals of care conversations, initiate advance care planning, provide patient education about palliative care, and care coordination related to their palliative care needs. The research nurses will work with the primary care provider (PCP) of each patient to implement the primary palliative care plan. For example, if the patient reports pain and needs any medications, the research nurse will first consult with the study palliative care physician and then work with the patient to craft a personalized care plan. If the patient and their caregiver want to contact their PCP, they could do so themselves. If the patient/caregiver wants the primary palliative care research nurse to contact the PCP on their behalf, the nurse will do so. All medical therapies will be prescribed by the individual patient’s primary care provider. These patients and their caregivers will also have email and a phone number to reach the team of primary palliative care research nurses in the study and they can contact them for any specific needs and questions during the intervention period.

### Procedure for handling any requests from patients in the usual care arm during the study period

Participants in the usual care arm will not be assigned to a primary palliative care research nurse. When participants in the usual care arm express any care needs to the study evaluators during evaluation meetings or if they contact the study team through phone or email, their requests will be routed to the study project manager. After contacting the patient, the study manager will acknowledge receipt of their request and recommend that the patient contact their PCP directly. If the patient requests it, the study manager will contact the patient’s care team as a courtesy and convey the patient’s request.

### Potential Risks

Primary palliative care is known to be beneficial and not risky. We highlight that all the medical treatments will be done by or through the individual patient’s primary care provider allowing for careful care coordination. The patient’s primary care provider will serve as the attending physician and sign the POLST form after discussion with the patients. As the patient’s caregiver will be involved in the primary palliative care plan, and as the individual patient’s primary care provider will be providing all medical therapies, we anticipate care coordination and improvement of the care quality at no risk to the patient.

Risks of answering the study survey instruments are minimal and may occasionally cause distress. In all cases the patient (and proxy) will have the right to refuse to answer any questions or drop out at any time. The project PI is a geriatrician and palliative care doctor who has worked with dementia patients for the past 15 years. She will work closely with the project staff to provide support to all patients enrolled. Any patient will be able to reach out to Dr. Periyakoil and also confidentially to the IRB for support (phone numbers are listed on the consent form).

### Recruitment and Informed Consent

After securing appropriate IRB permission, we will work with the neurology team to identify eligible patients for our study. The research staff will explain the consent form and what participation in the study involves.

### Protection from risks

In an effort to mitigate any risks to the subjects, the study design was created by Dr. Periyakoil (PI) and Dr. Helena Kraemer (Chief Biostatistician) with input Drs.Von Gunten and Covinsky. Dr. Frank Ferris serves as the safety monitor and provides oversight of the data safety and management.

### Planned procedures for protecting against assessment complications

There are few if any risks in completing the study intervention (usual care enhanced by early primary palliative care).

### Monitoring Procedures and Treatment for Adverse Reactions

The project team will meet weekly to review every aspect of the study and discuss any adverse reactions. Any adverse reactions will be reported to Stanford IRB as per our standard protocol. At any point in the protocol a treatment failure may be declared and the patient has the option of opting out of the intervention at any time during the study. Patients are free to withdraw their consent and to discontinue participation at any time without prejudice to them or effect on their (patients’) medical care.

### Planned procedures for protection of confidentiality

All data will be coded by subject number to ensure anonymity of participation. Data will be kept in locked cabinets and secured computer files. Online data will be hosted on a secure server at Stanford University and available through double authorization protocol for maximal security. Research records will be kept confidential to the level allowed by law. The findings of this research may be published; however, neither the name nor identity of the subjects or their caregivers will be used in any publications. Only the study manager and database manager will have access to the research database.

## POTENTIAL BENEFITS OF THE PROPOSED RESEARCH TO THE SUBJECTS AND TO OTHERS

*Overview:* It is expected that the risk vs. benefits ratio of this project will be favorable. At worst, patients can drop out and receive usual care. All study participants will be provided with a courtesy stipend for their annual participation in the study outcome measures. We anticipate that these patients will benefit from the early primary palliative care intervention. Beyond the benefit for the individual patient, this study has a transformational potential as it can identify the common primary palliative care needs of patients with Mild Cognitive Impairment (MCI), mild and moderate dementia living in the community as well as their caregivers. If successful, it is a scalable, low risk solution that is patient-centered and family-oriented with minimal risks to participants and its benefits of the proposed project far outweigh any risks. Once we complete the study, we will be able to refine the intervention based on our findings and test in real world environments.

## STATISTICAL ANALYSIS

Each of the primary outcome measures are measured repeated over time, at baseline, months 4 and 12.

To compare the early palliative care intervention (EPC) and usual care (UC) groups, for ordinal outcome measures, we will use a Hierarchical Linear Model (HLM) that assumes that the Outcome (O) for each patient (p) at time (t) is a linear function with an intercept (i) and slope (s): O(p,t)=i(p)+s(p)ln(t+1)+e (t,p), where t is the time of measurement (measured from baseline), and e is an error term. The intercept for each patient (i(p)) is the true baseline response, with randomization having the same distribution in both groups. The slopes in the groups reflect the impact of treatment (EPC or UC) over the course of treatment. Both intercepts and slopes are assumed to have a bivariate distribution in the population of participants, and the error terms within participants are assumed to be intercorrelated according to an autoregressive covariance function (outcomes closer together assumed to be more highly correlated that those more distal to each other). Also, ln(t+1) is used rather than t, because outcomes in randomized clinical trials are more likely to have a “fishhook” relationship to time (t), with more rapid changes early on than later91. In this intervention this is even more likely since the intervention is more intensive in year 1 than later.

**Power calculation:** The null hypothesis to be tested is that the slopes in the EPC group have the same distribution as those in the UC group, i.e., that there is no effect of EPC relative to UC. The significance level is to be 5%, with no adjustment for multiple testing because the outcomes chosen tend to be independent of each other, a two-tailed test to be conservative. For 80% power to detect a moderate effect size with a two-tailed 5% test, the sample size per group is approximately 63. However, we propose to recruit a total of 100 participants per arm (200 in total) as we anticipate persons with CRD=0 and at risk of cognitive impairment and those with CDR=0.5,1 or 2 and we can complete within group analysis.

## IMPORTANCE OF KNOWLEDGE TO BE GAINED

Currently very little is known about the primary palliative needs of patients with cognitive impairment living in the community. If successful, this trial will delineate the common primary palliative care needs of patients at risk of cognitive impairment and those with MCI, mild and moderate dementia. Such knowledge will allow for future interventions to better support persons living with dementia in the community.

## SUPPLEMENTAL MEASURES

### EVALUATION MEASURES FOR ALL STUDY PARTICIPANTS

These measures were administered to patients in both the intervention arm and the usual care arm by evaluators who were blinded to the treatment conditions) at Day 0, 4 months, and one year. Ongoing longitudinal, observational follow up will be conducted to trace study patients over time to better understand the durability of the ELICIT intervention on the care of the participants. For every study participant in both arms, the study manager tracked the number of supportive care needs expressed by the patients, their completion dates of the WMM and the POLST in the medical records.

### Edmonton Symptom Assessment Scale(ESAS)[10]

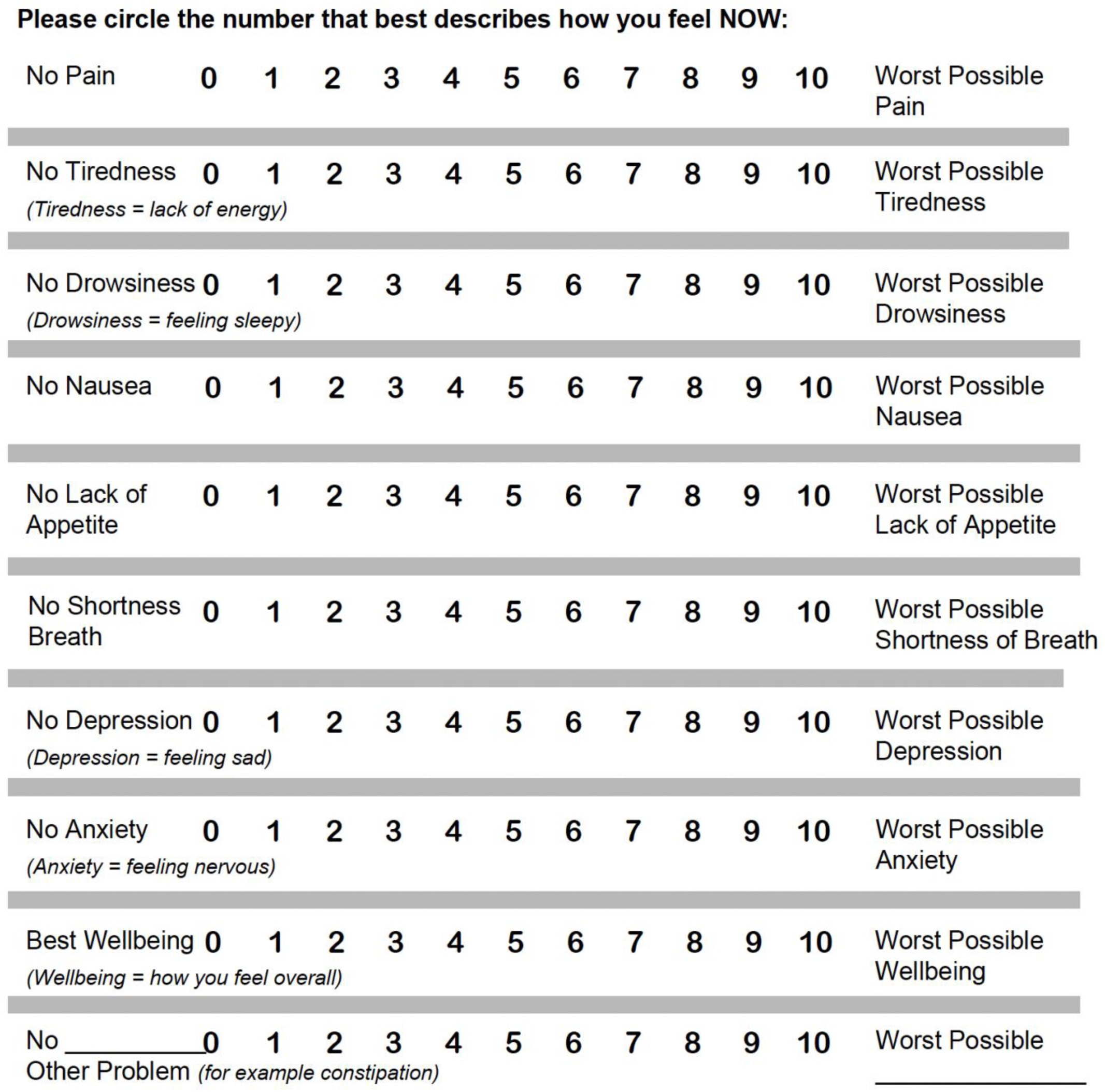

### Quality of Life: Alzheimer’s Disease measure(QoL-AD)[11]

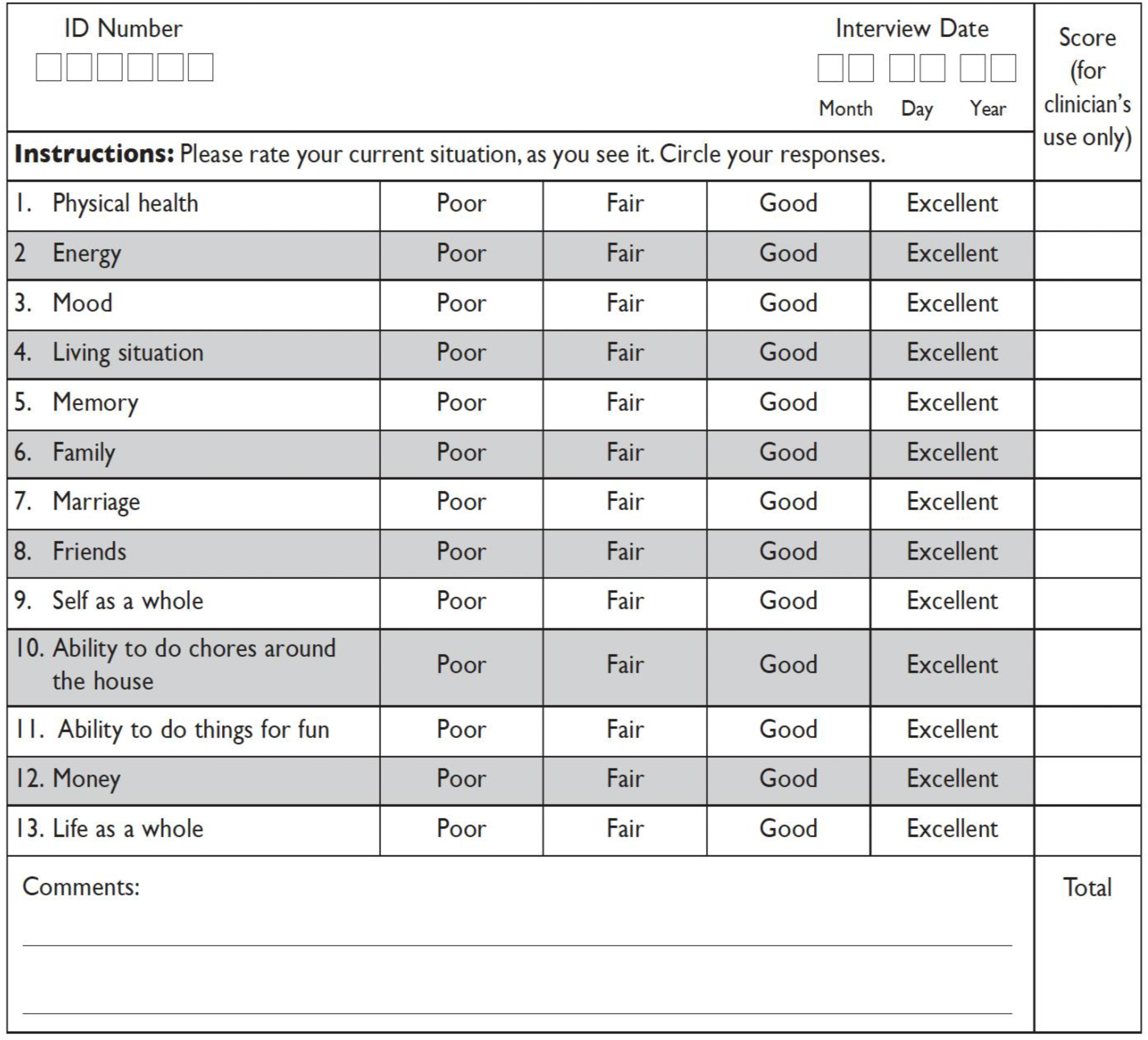

### Patient Activation Measure (PAM)[12]

Below are some statements that people sometimes make when they talk about their health. Please indicate how much you agree or disagree with each statement as it applies to you personally by circling your answer. There are no right or wrong answers, just what is true for you. If the statement does not apply to you, circle N/A.

|  |  |  |  |  |  |
| --- | --- | --- | --- | --- | --- |
| 1. I am the person who is responsible for taking care of my health. | Disagree Strongly | Disagree | Agree | Agree Strongly | N/A |
| 2. Taking an active role in my own health care is the most important thing that affects my health. | Disagree Strongly | Disagree | Agree | Agree Strongly | N/A |
| 3. I am confident I can help prevent or reduce problems associated with my health. | Disagree Strongly | Disagree | Agree | Agree Strongly | N/A |
| 4. I know what each of my prescribed medications do. | Disagree Strongly | Disagree | Agree | Agree Strongly | N/A |
| 5. I am confident that I can tell whether I need to go to the doctor or whether I can take care of a health problem myself. | Disagree Strongly | Disagree | Agree | Agree Strongly | N/A |
| 6. I am confident that I can tell a doctor or nurse concerns I have even when he or she does not ask. | Disagree Strongly | Disagree | Agree | Agree Strongly | N/A |
| 7. I am confident that I can carry out medical treatments I may need to do at home. | Disagree Strongly | Disagree | Agree | Agree Strongly | N/A |
| 8. I understand my health problems and what causes them. | Disagree Strongly | Disagree | Agree | Agree Strongly | N/A |
| 9. I know what treatments are available for my health problems. | Disagree Strongly | Disagree | Agree | Agree Strongly | N/A |
| 10. I have been able to maintain lifestyle changes, like healthy eating or exercising. | Disagree Strongly | Disagree | Agree | Agree Strongly | N/A |
| 11. I know how to prevent problems with my health. | Disagree Strongly | Disagree | Agree | Agree Strongly | N/A |
| 12. I am confident I can work out solutions when new problems arise with my health. | Disagree Strongly | Disagree | Agree | Agree Strongly | N/A |
| 13. I am confident that I can maintain lifestyle changes, like healthy eating and exercising, even during times of stress. | Disagree Strongly | Disagree | Agree | Agree Strongly | N/A |

## MEASURES USED BY THE NURSES FOR PATIENTS IN THE PRIMARY PALLIATIVE CARE INTERVENTION ARM

This virtual, nurse-led intervention focused on the four domains of primary palliative care: (a) assessing and palliating physical symptoms, (b) assessing and managing psychological, social, cultural, and spiritual aspects of care, (c) establishing goals of care, identification of a surrogate decision maker and completion of advance care planning documentation and (d) providing care coordination support.

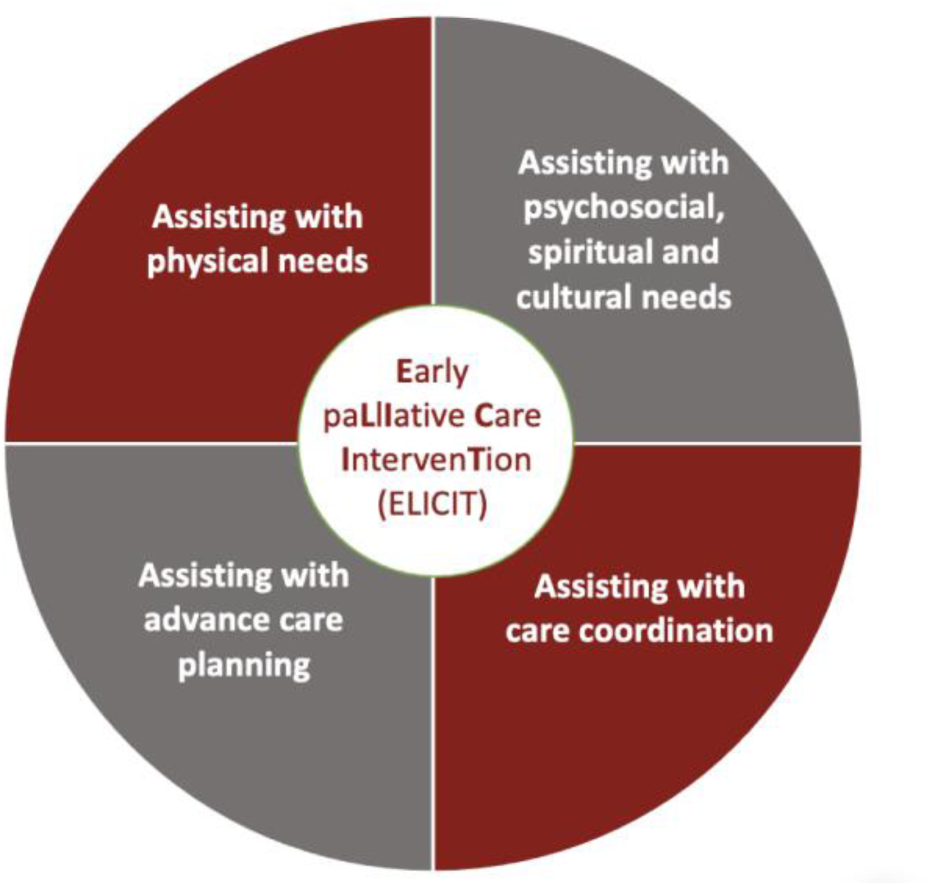

### Distress thermometer[13]

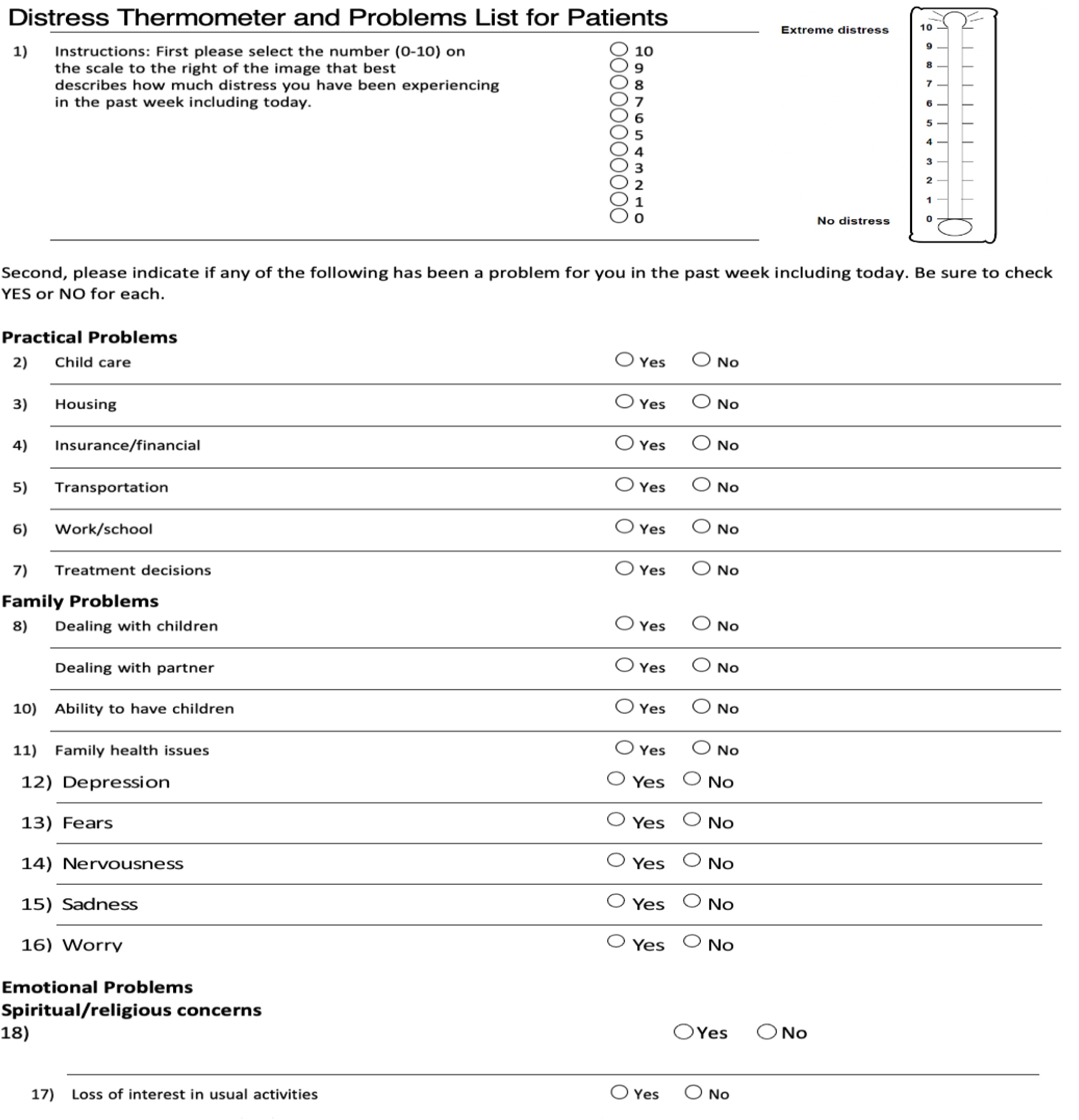
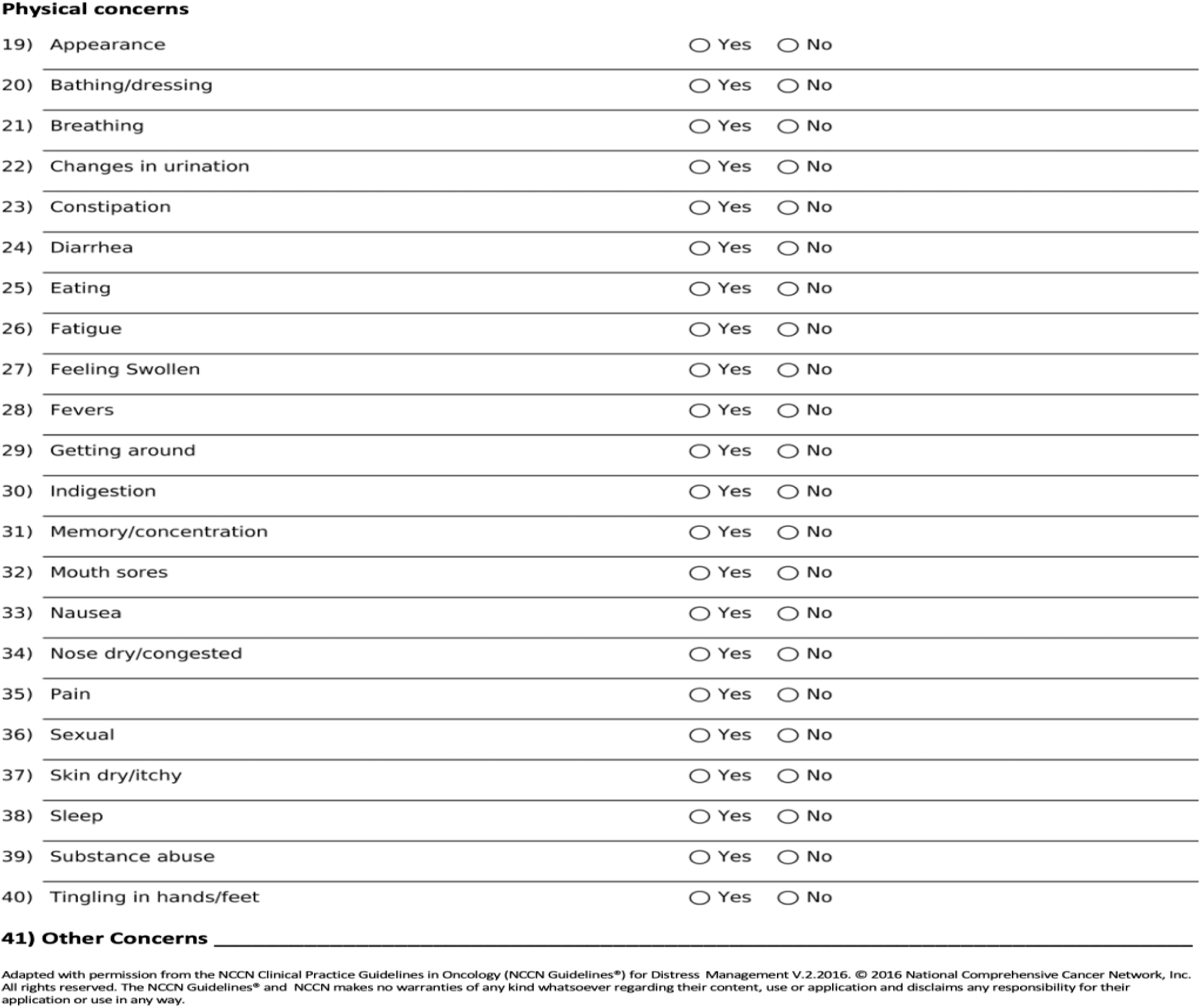

### KATZ ADLs scale[14]

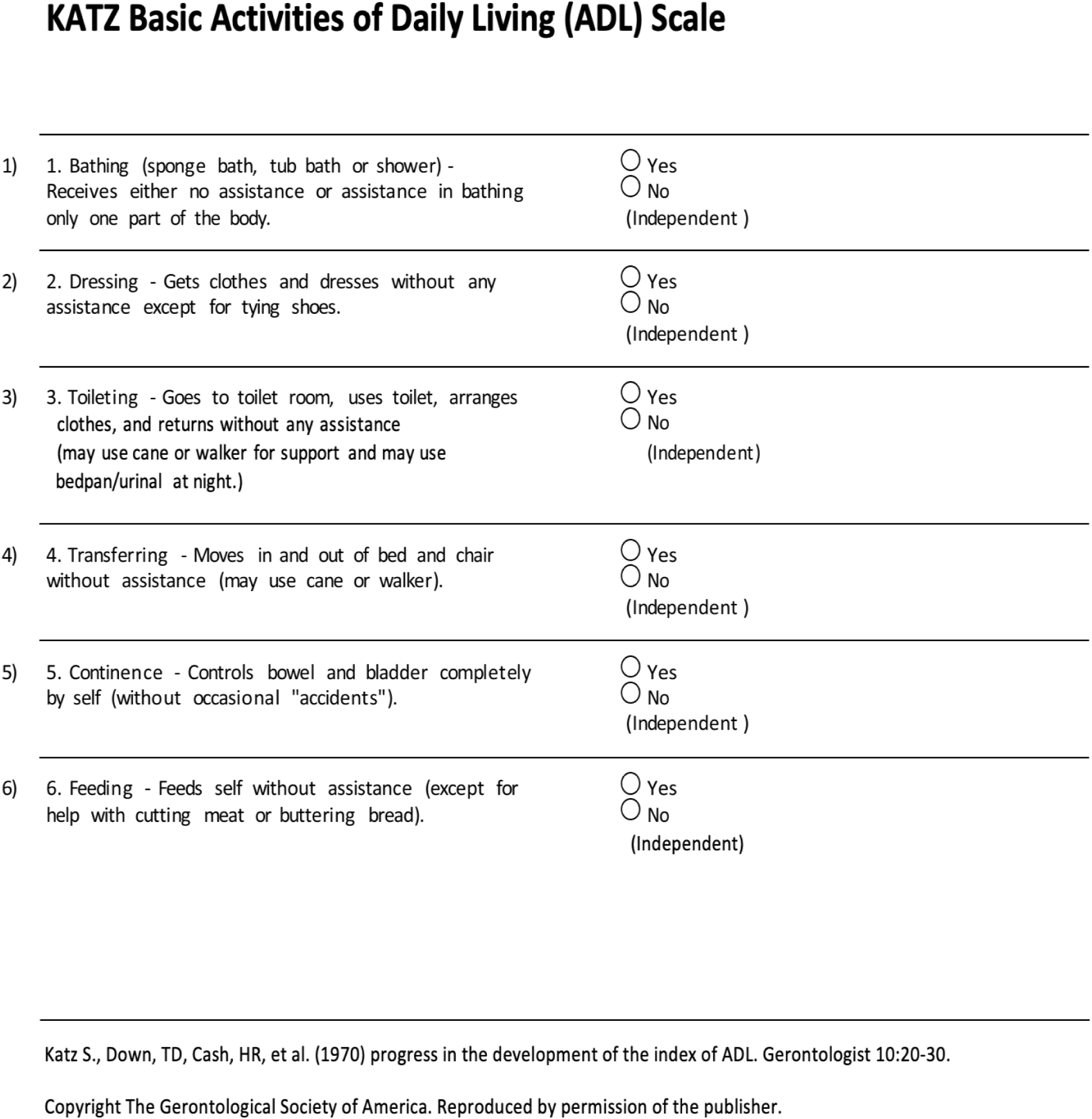

### Spiritual Assessment Exploratory Questions

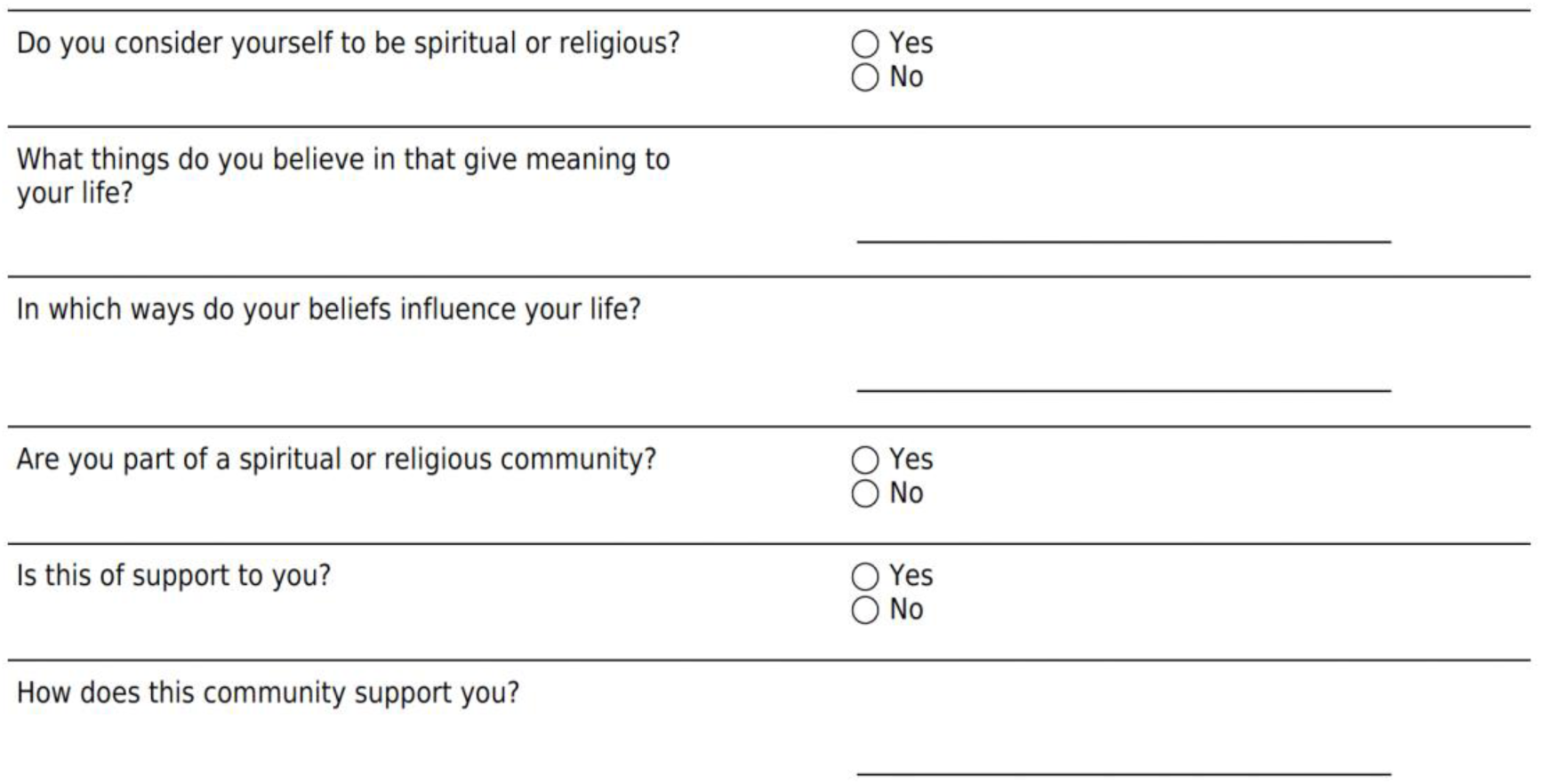

### Social Assessment Exploratory Questions

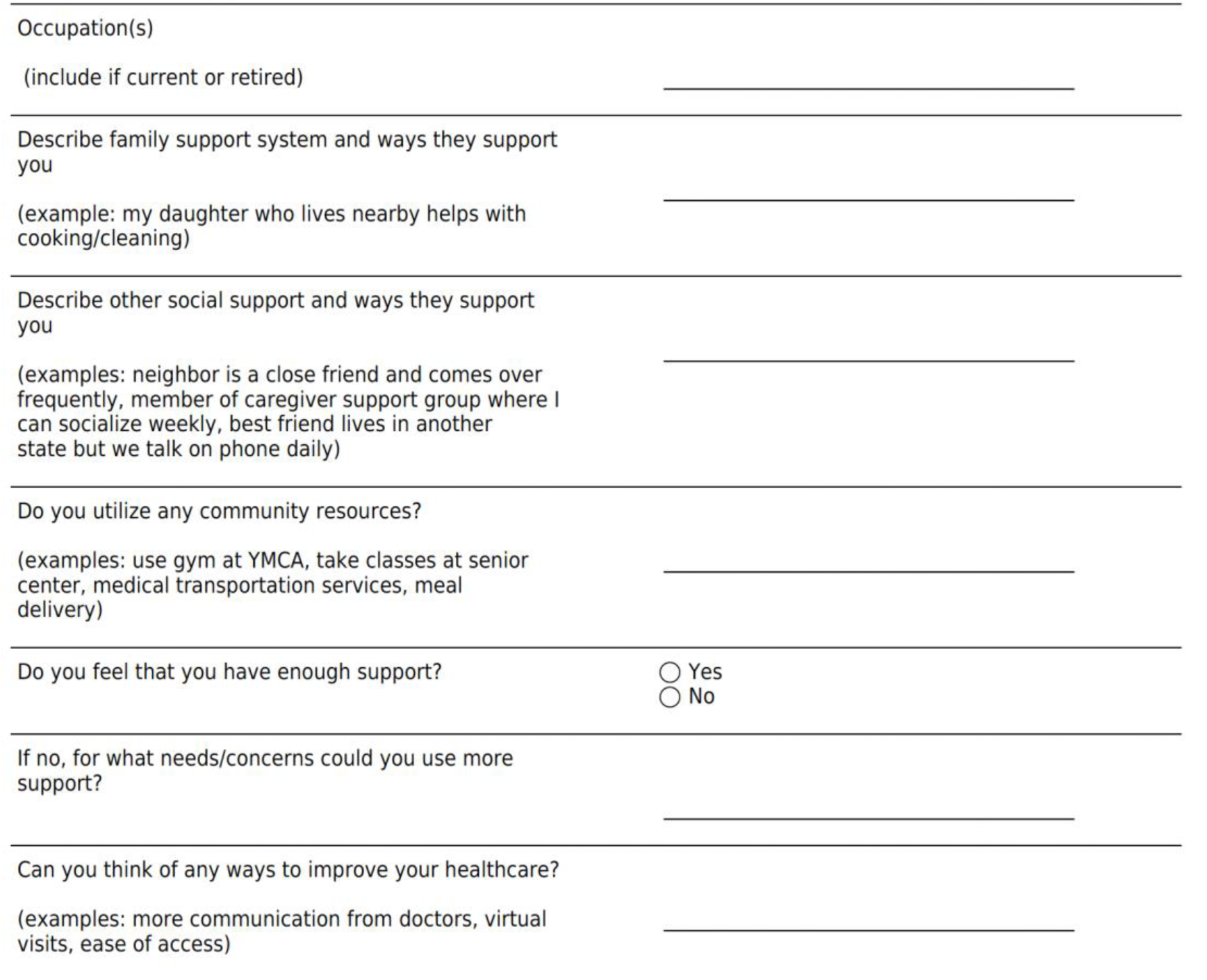

### Pain Assessment

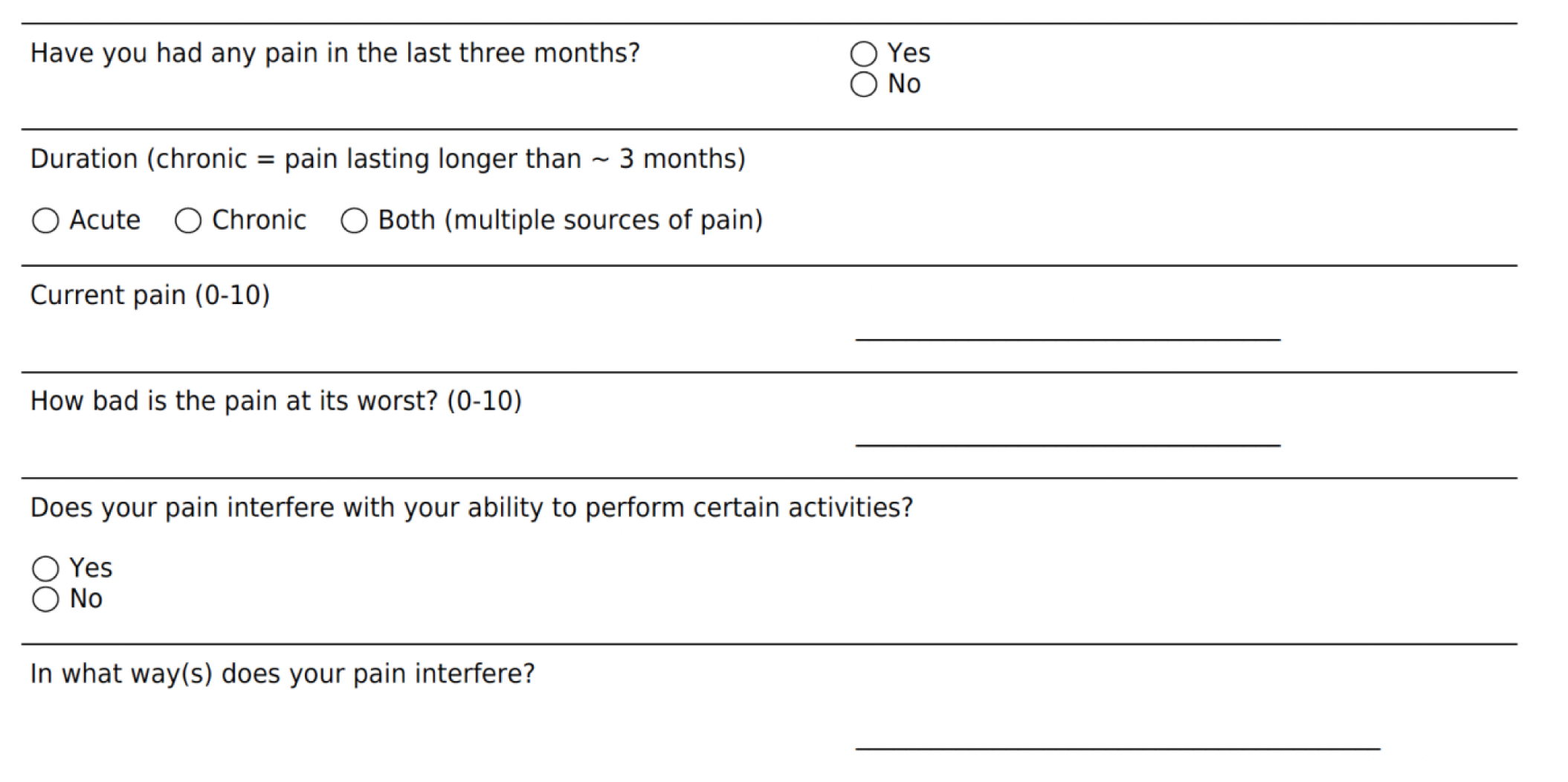

### Pain Location

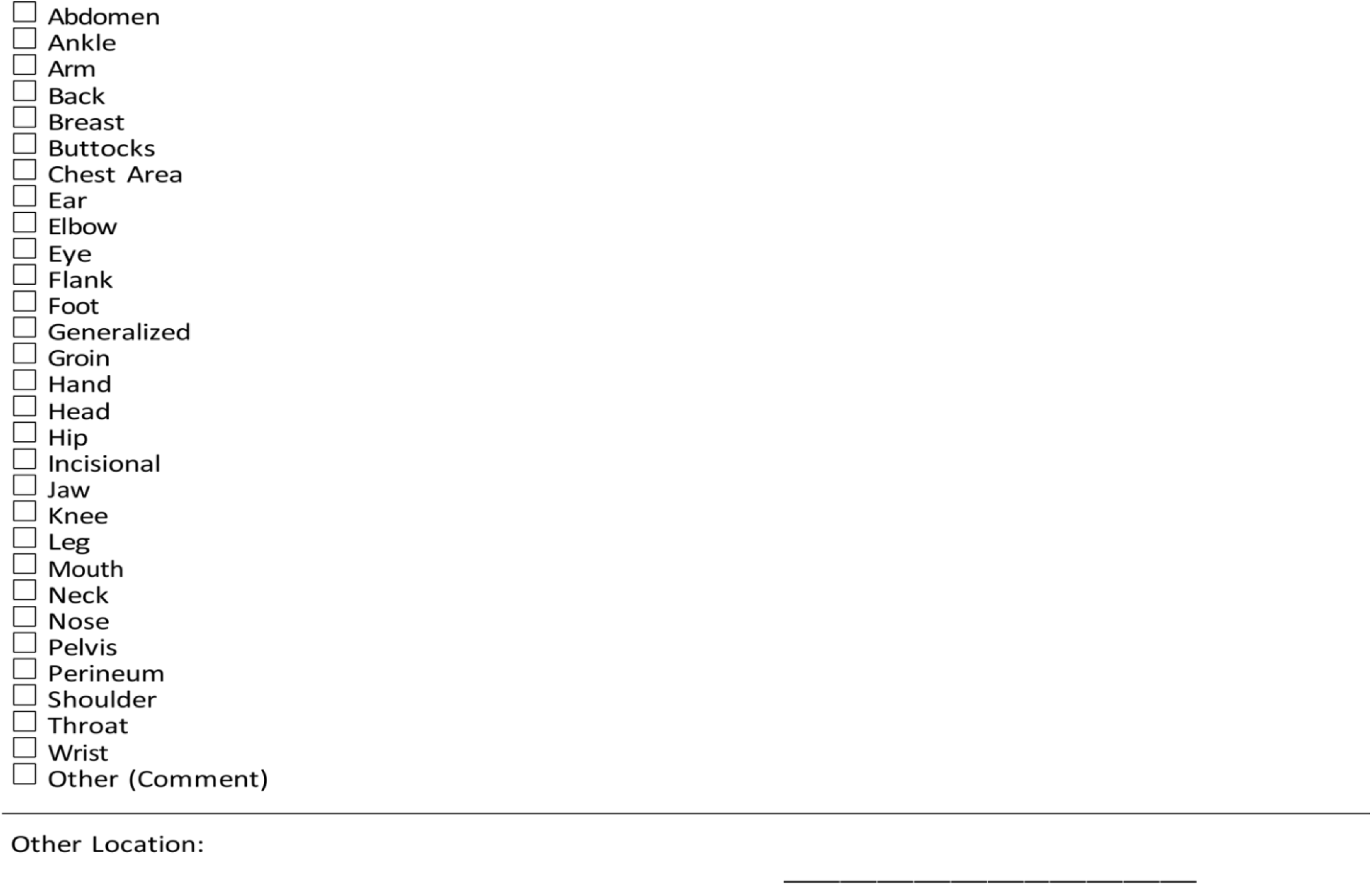

### Description

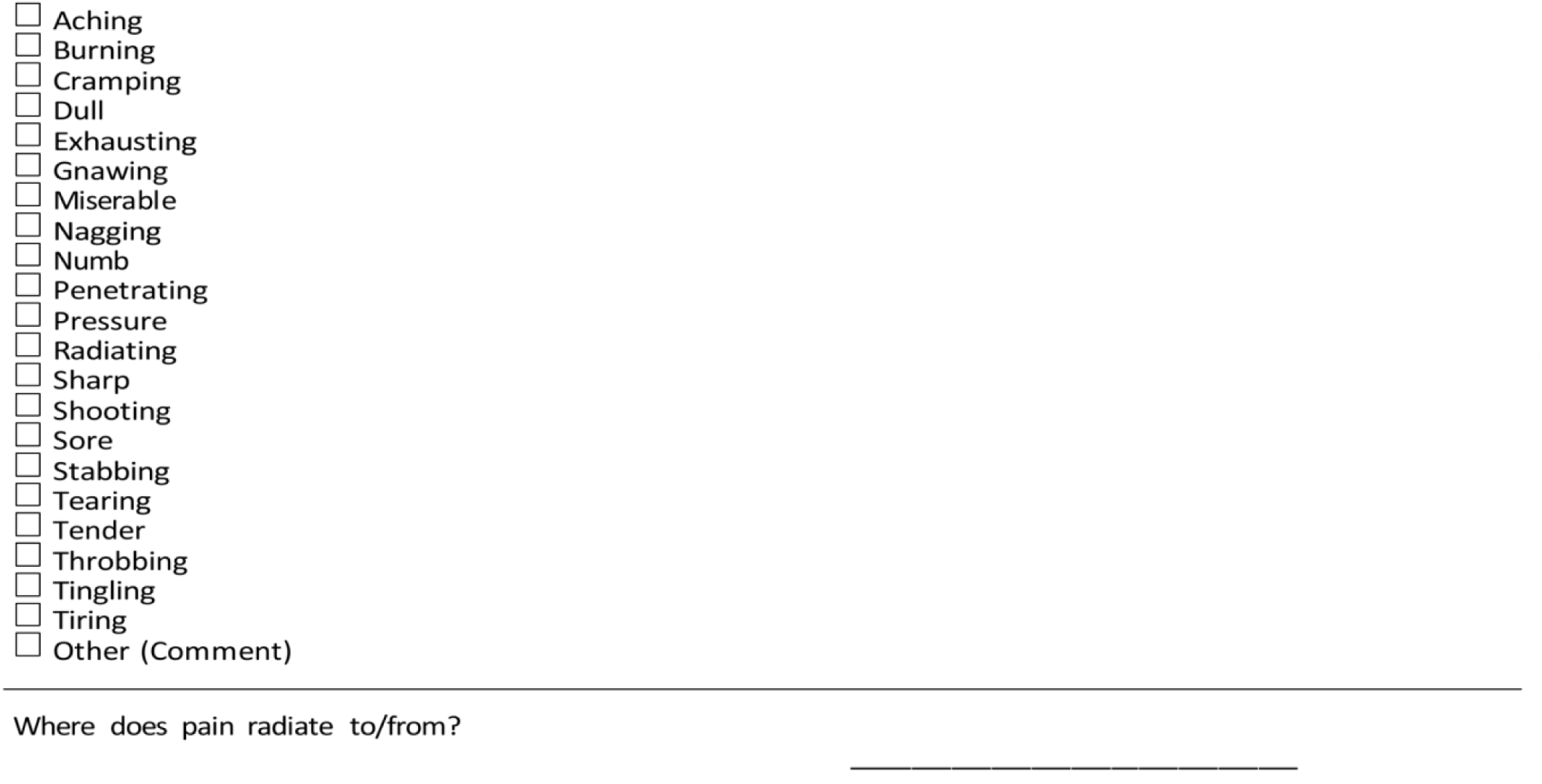

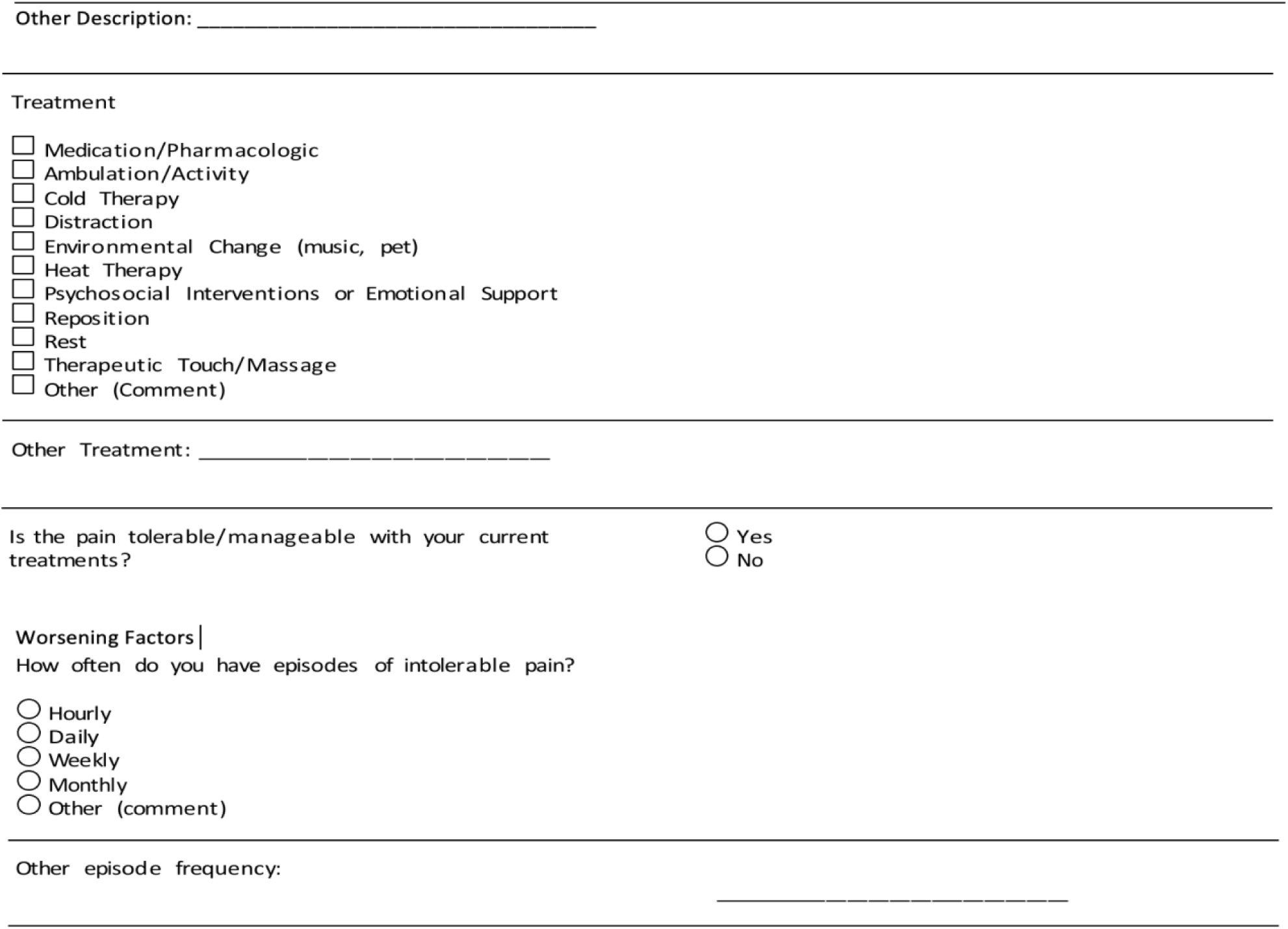

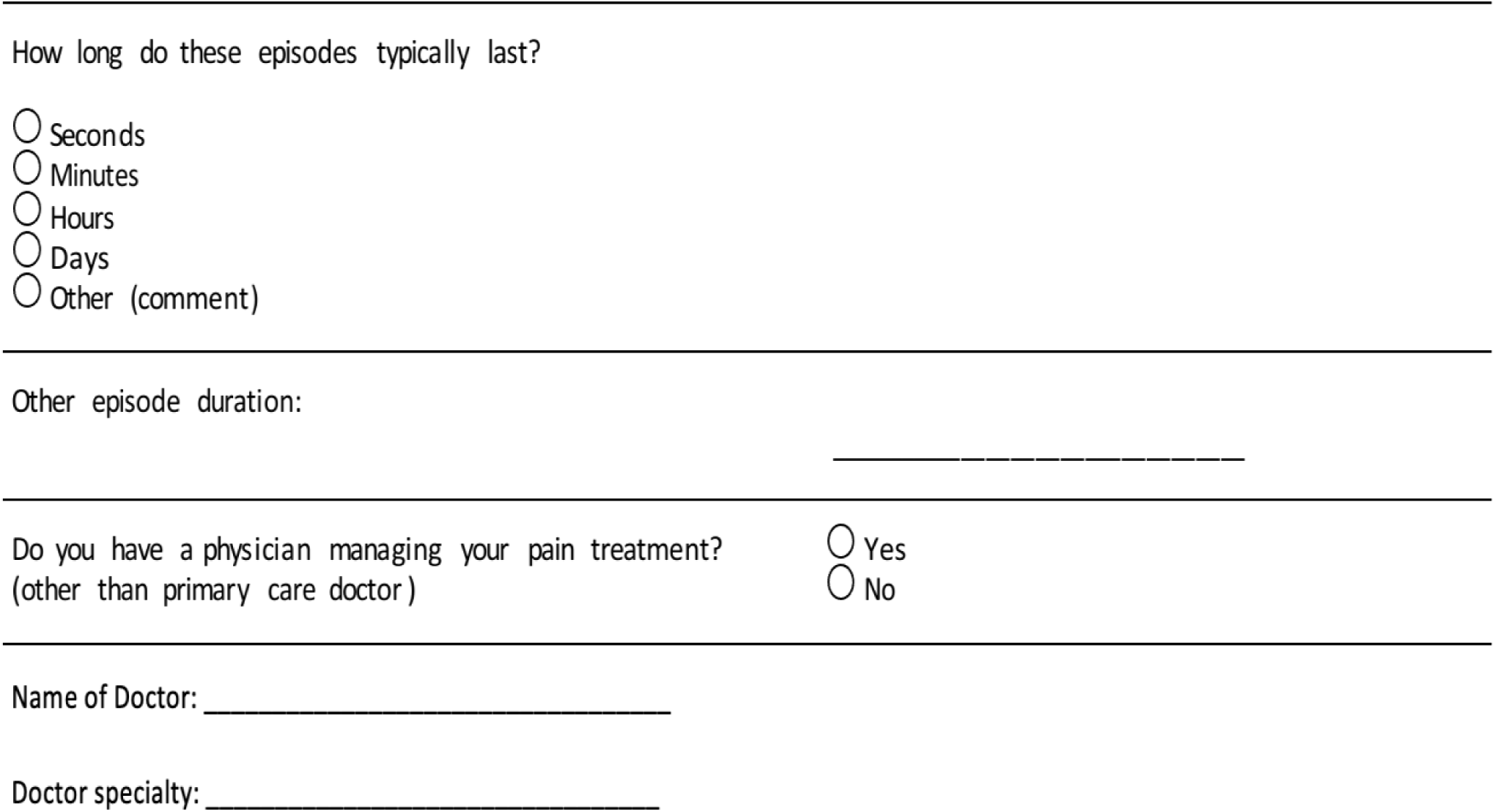

Any other notes based on assessment:

### Depression assessment using the Geriatric Depression Scale [15]

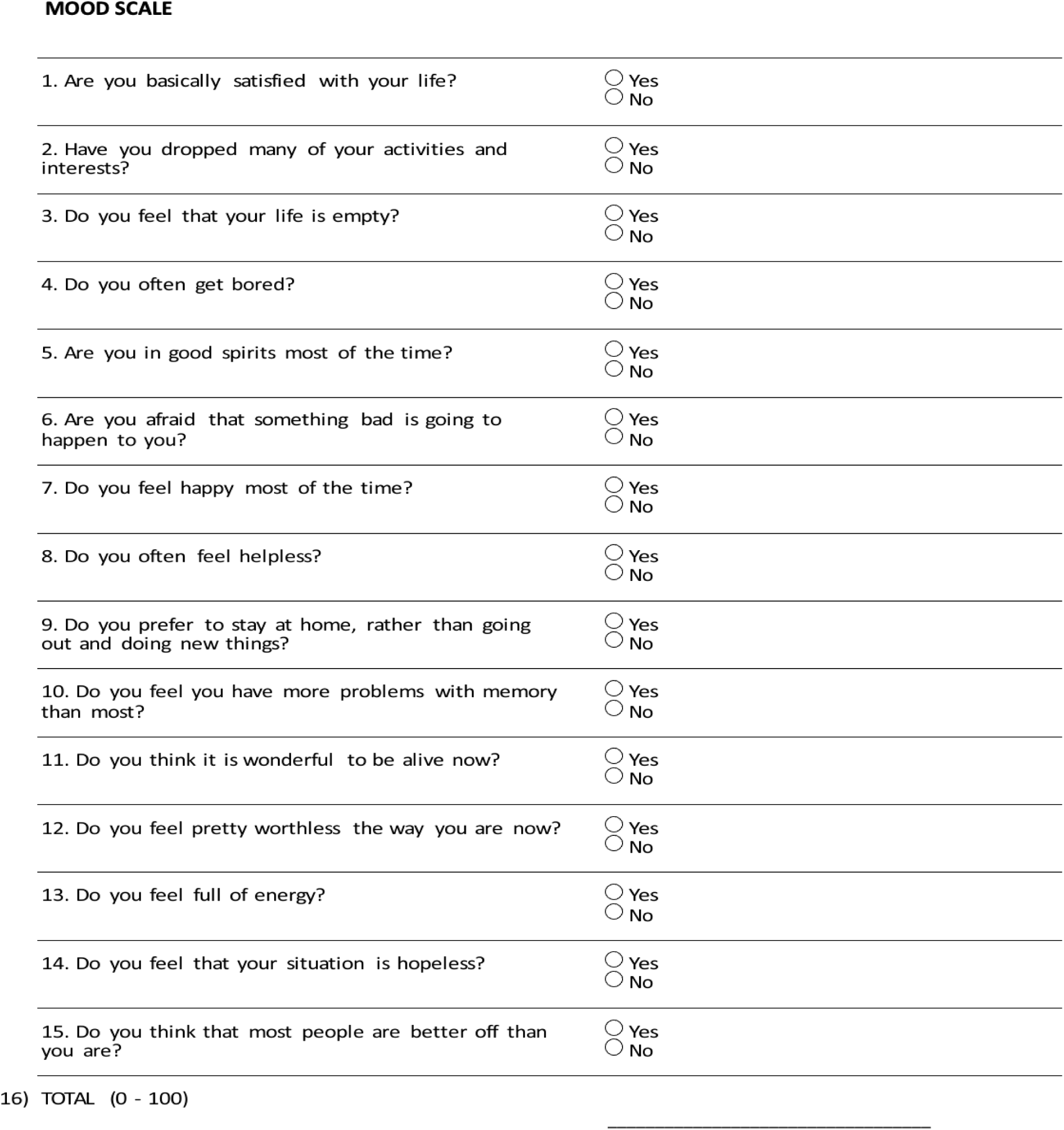

### Life Goal List[16]

Aten steps approach to writing a Life Goal List:

1. Reflect on what matters most to you and your core values.
2. Next identify some of the life experiences you would like to have or tasks you would like to complete
3. Identify the timeline for when you want to complete each item on your life goal list. Some may take years and others can be achieved relatively soon.
4. Use the table below to brainstorm a list of items for your life goal list.
5. Review your draft life goal list and trim it down. We recommend that you limit yourself to 4 to 5 items on your list. At least two of the life goals should be easy to achieve and in a short time. These easy wins will motivate you towards investing sustained effort towards more lofty goals.
6. Discuss your life goal list with your loved ones. They will often want to help you or even join you in your quests.
7. Talk to your doctor about your life goal list. Ask your doctor to personalize your medical care so you can accomplish the items on your life goal list.
8. Remember to check off at least one or two items on your life goal list every year.
9. Every time you check off a b life goal list item, pause for a moment and savor the sweet sense of a job well done.
10. On your birthday or another memorable day, be sure to review your life goal list and update it.

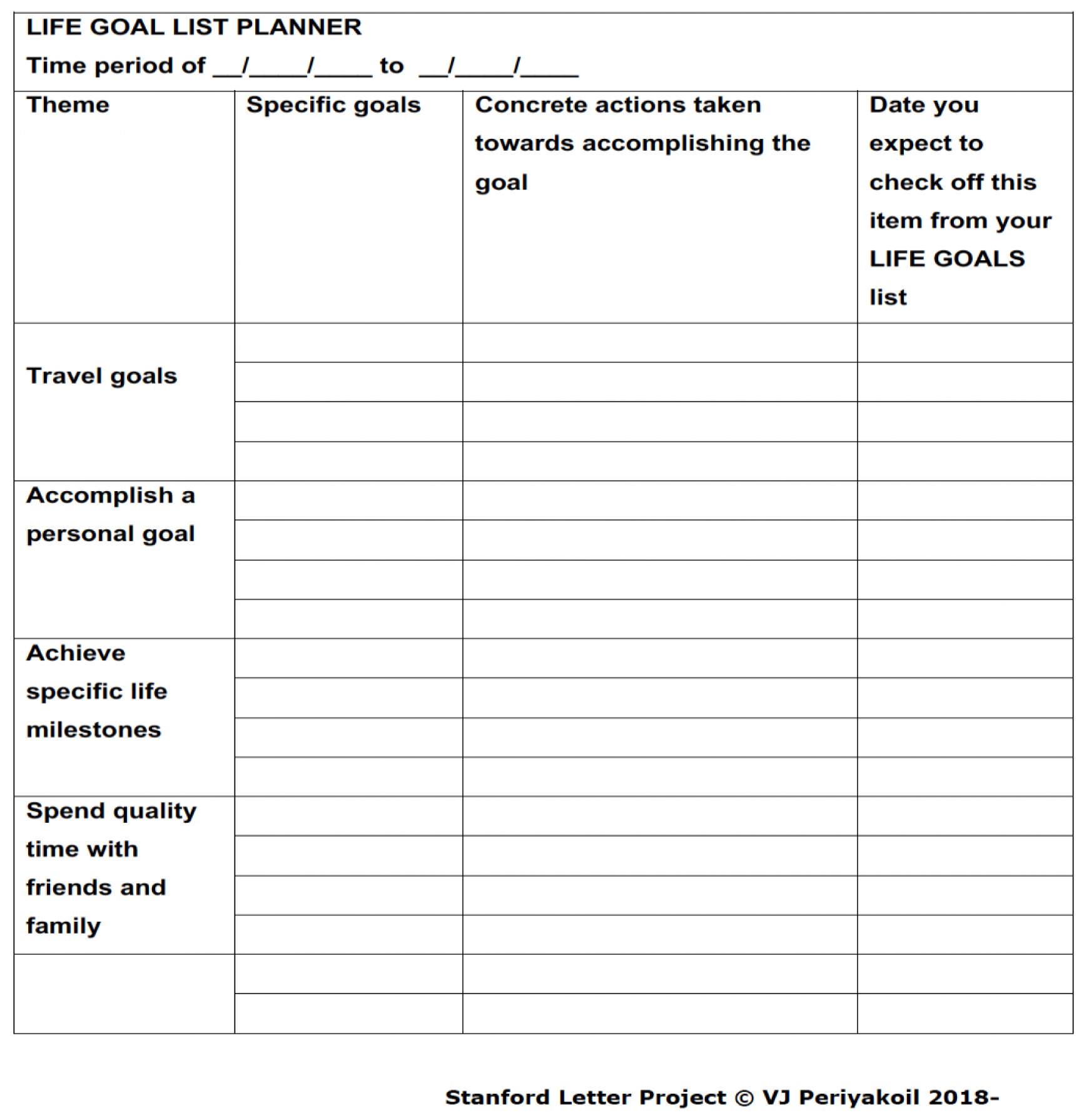
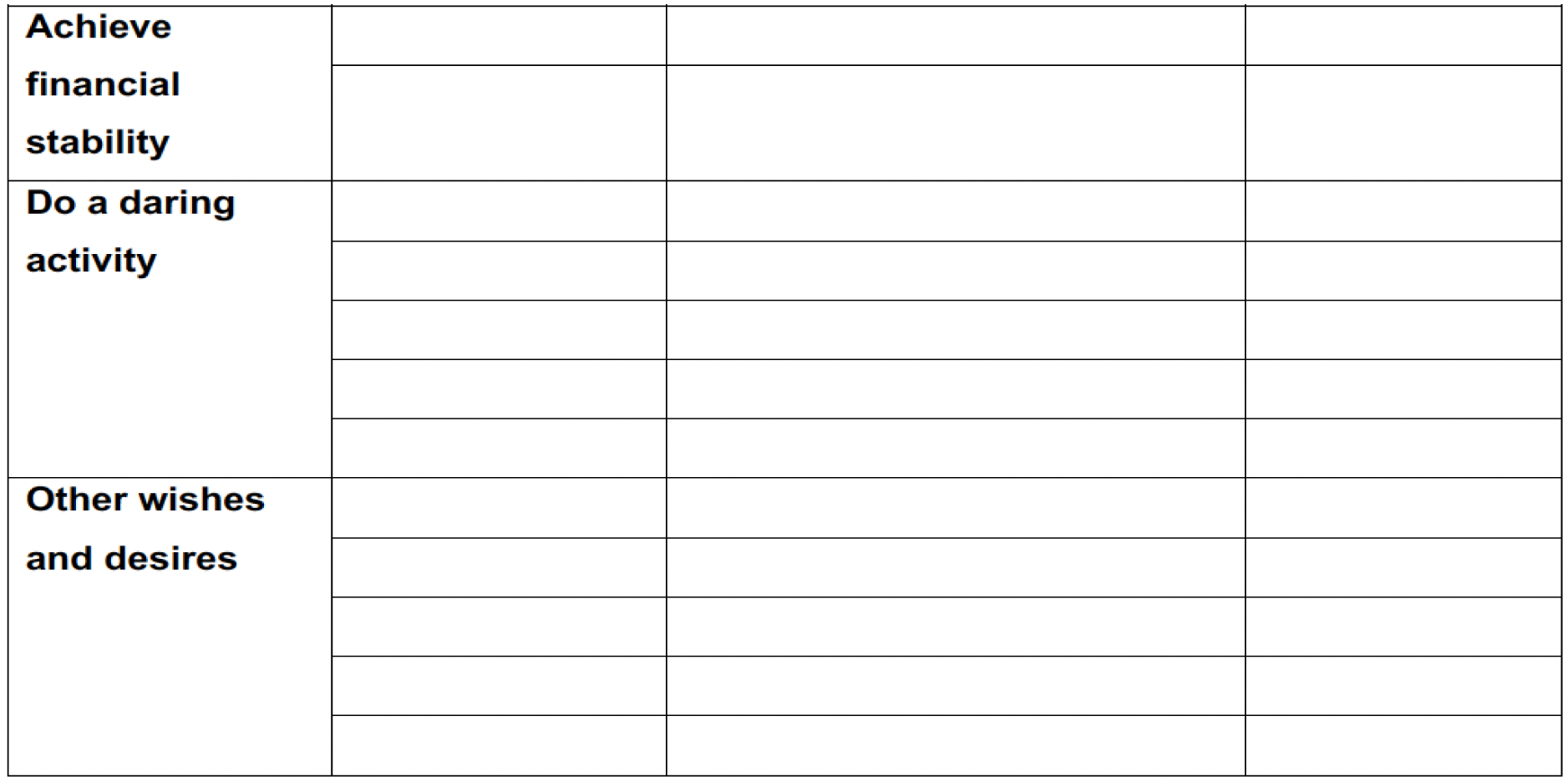

| LIFE GOAL LIST PLANNER |  |  |  |
| --- | --- | --- | --- |
| Time period of __/__/__ to __/__/__ |  |  |  |
| Theme | Specific goals | Concrete actions taken towards accomplishing the goal | Date you expect to check off this item from your LIFE GOALS list |
| Travel goals |  |  |  |
| Accomplish a personal goal |  |  |  |
| Achieve specific life milestones |  |  |  |
| Spend quality time with friends and family |  |  |  |
| **Achieve financial stability** |  |  |  |
| **Do a daring activity** |  |  |  |
| **Other wishes and desires** |  |  |  |

Review the table carefully and list 4 to 5 items in your LIFE GOAL list below

| LIFE GOAL LIST PLANNER |  |  |
| --- | --- | --- |
| Time period of __/__/__ to __/__/__ |  |  |
| Specific LIFE GOALS | Concrete actions to be taken towards accomplishing the goal | Anticipated date of completion |
| 1. |  |  |
| 2. |  |  |
| 3. |  |  |
| 4. |  |  |
| 5. |  |  |

### Physicians Orders for Life Sustaining Treatment (POLST)[17]

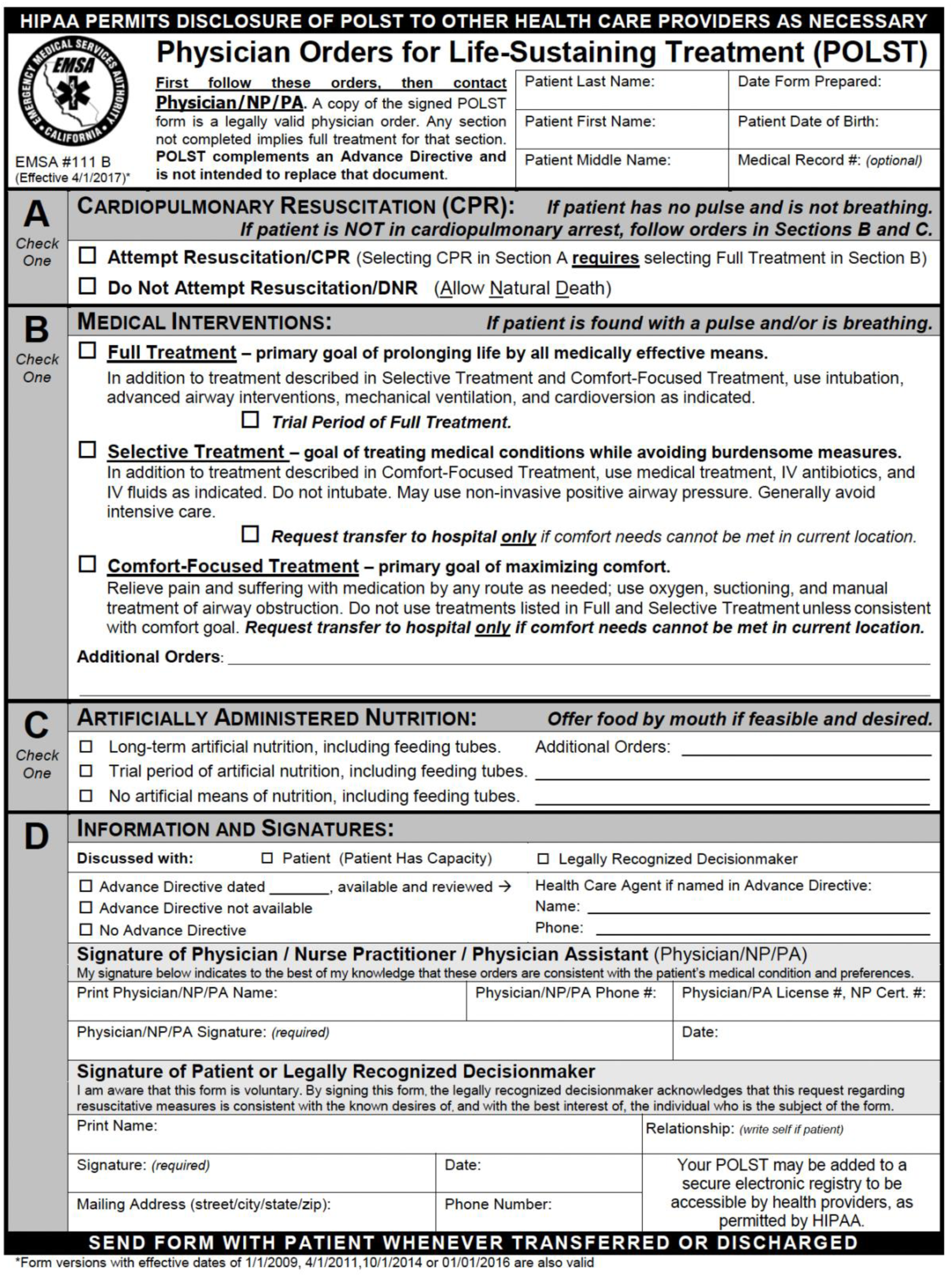

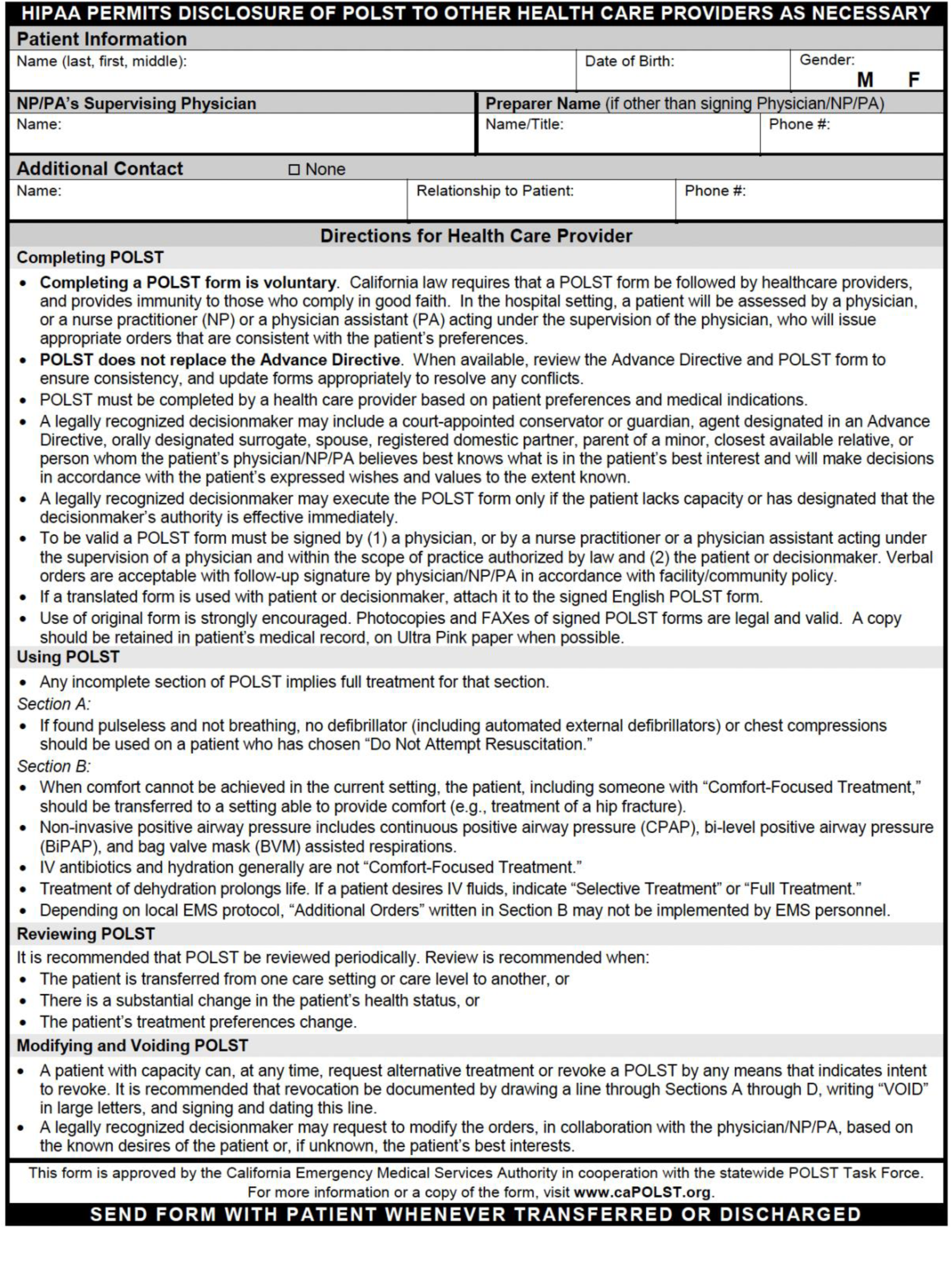

### What Matters Most Letter Advance Directive[18]

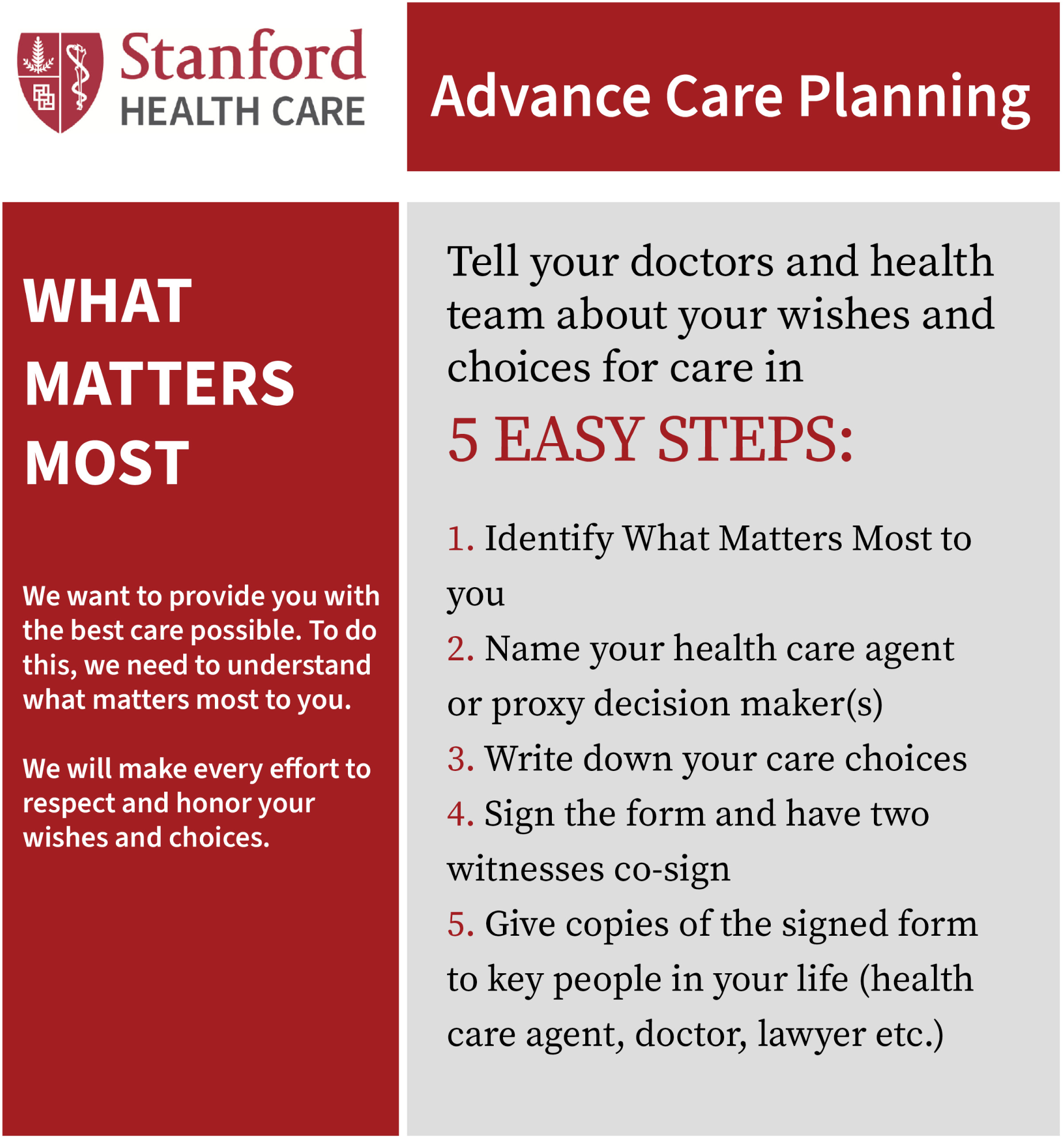

### Part 1: Tell Us about What Matters Most to You

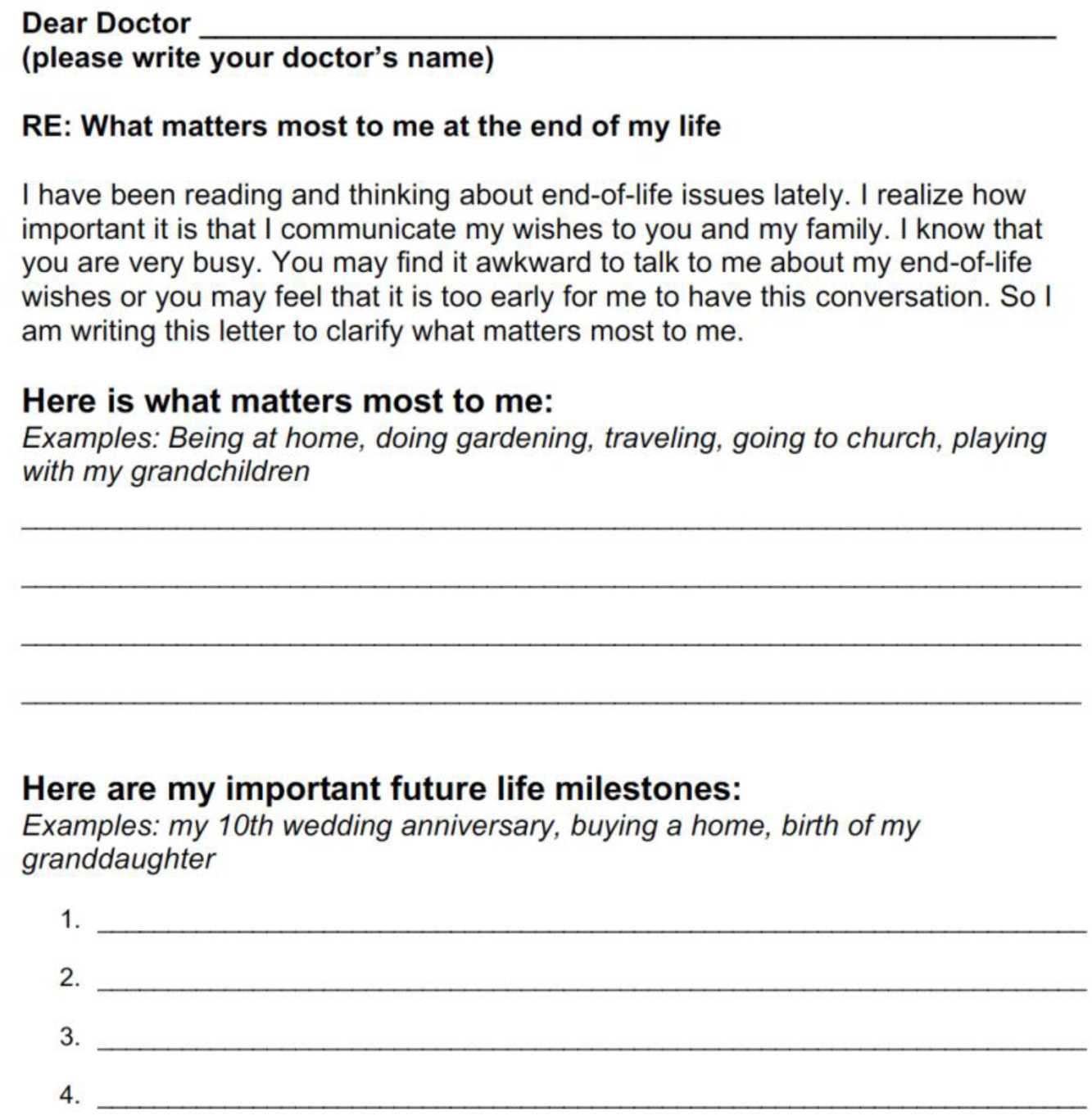
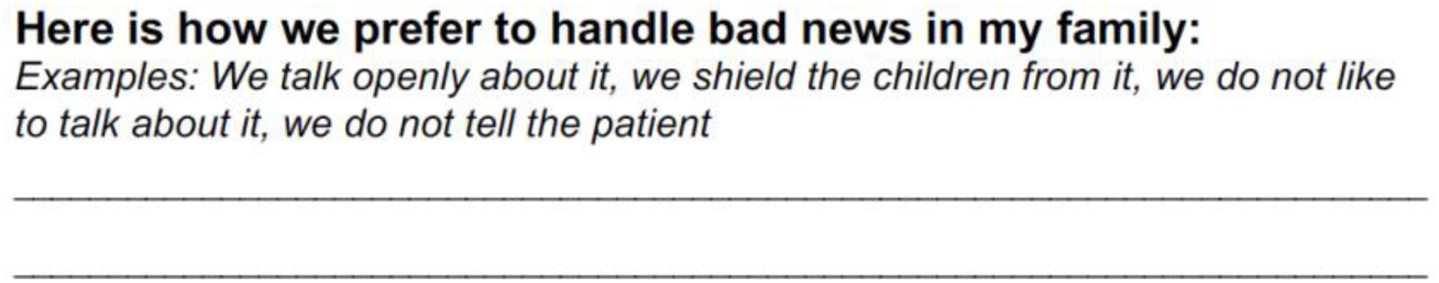

### Part 2: Who Makes Decisions for You when You Cannot

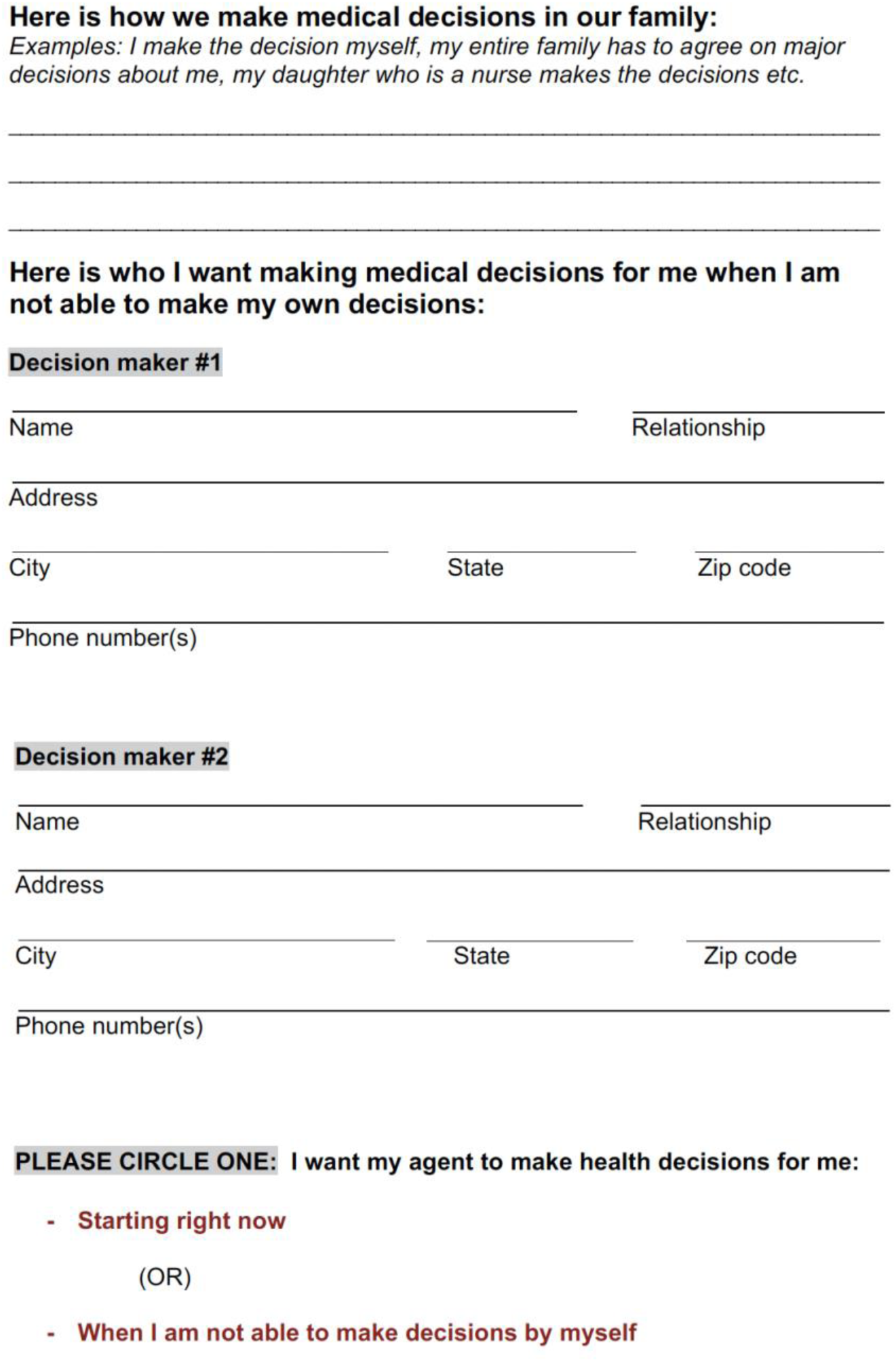

### Part 3: Please Write Down Your Care Choices

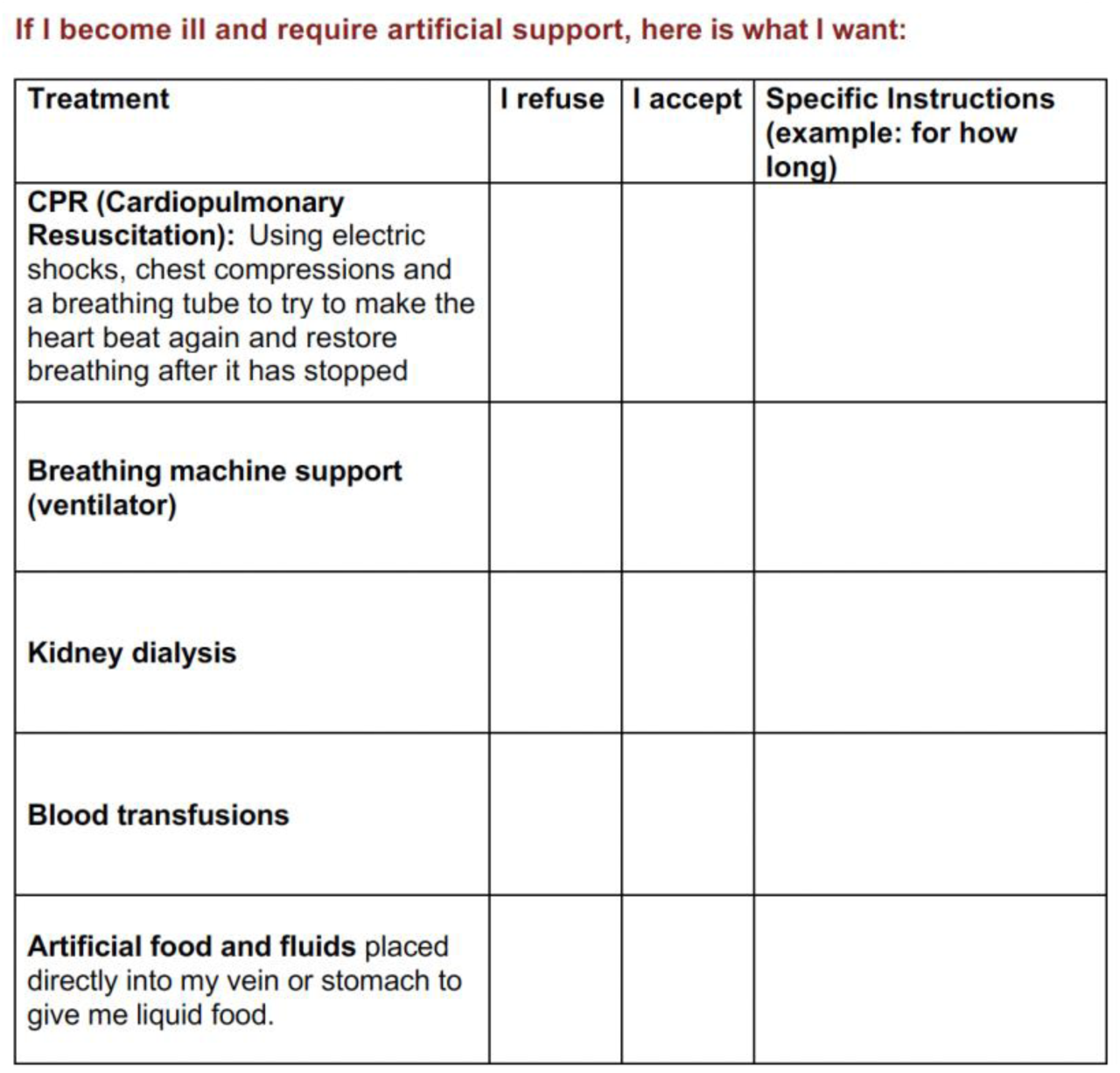

**Please allow natural death when**

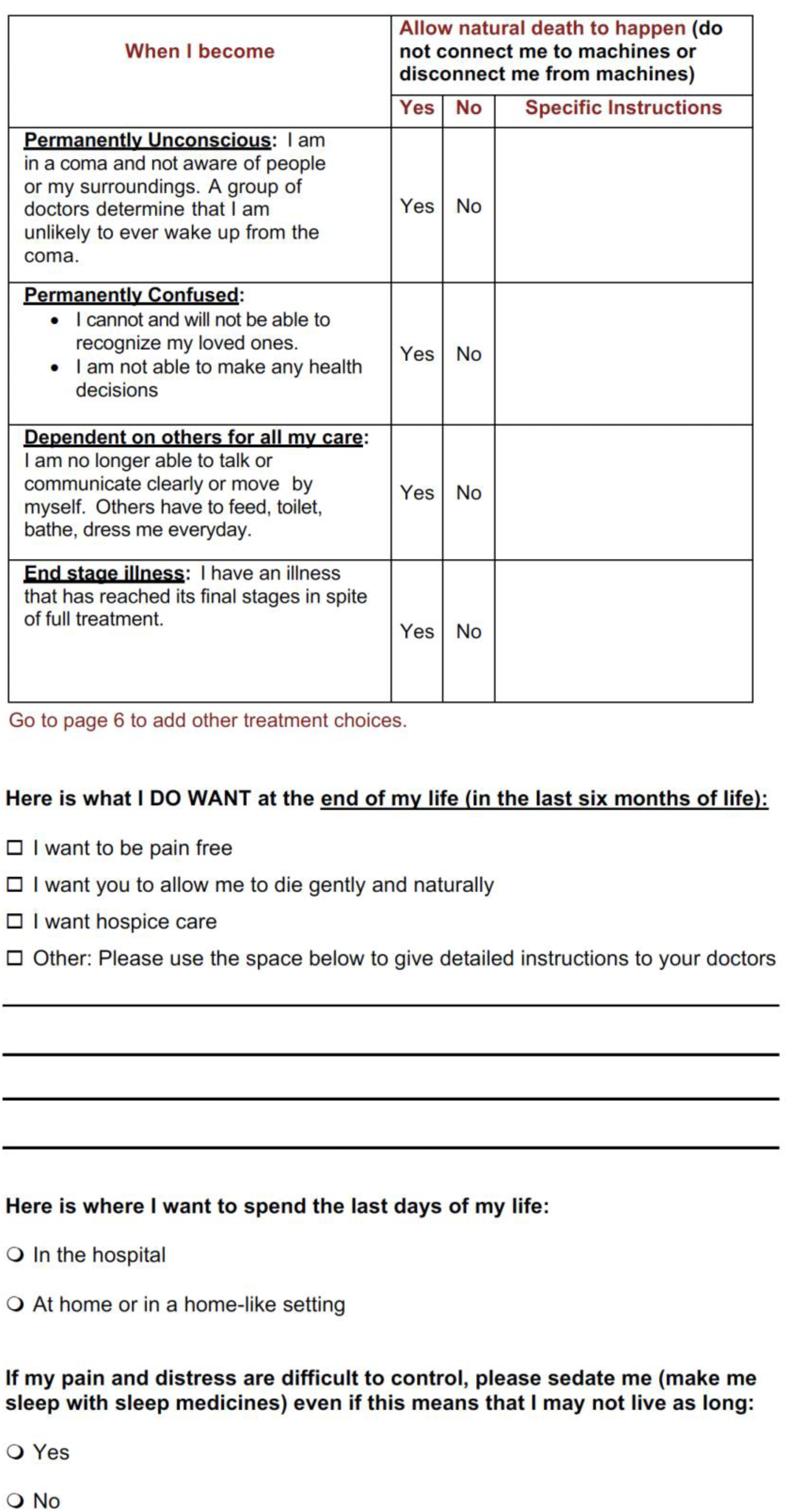
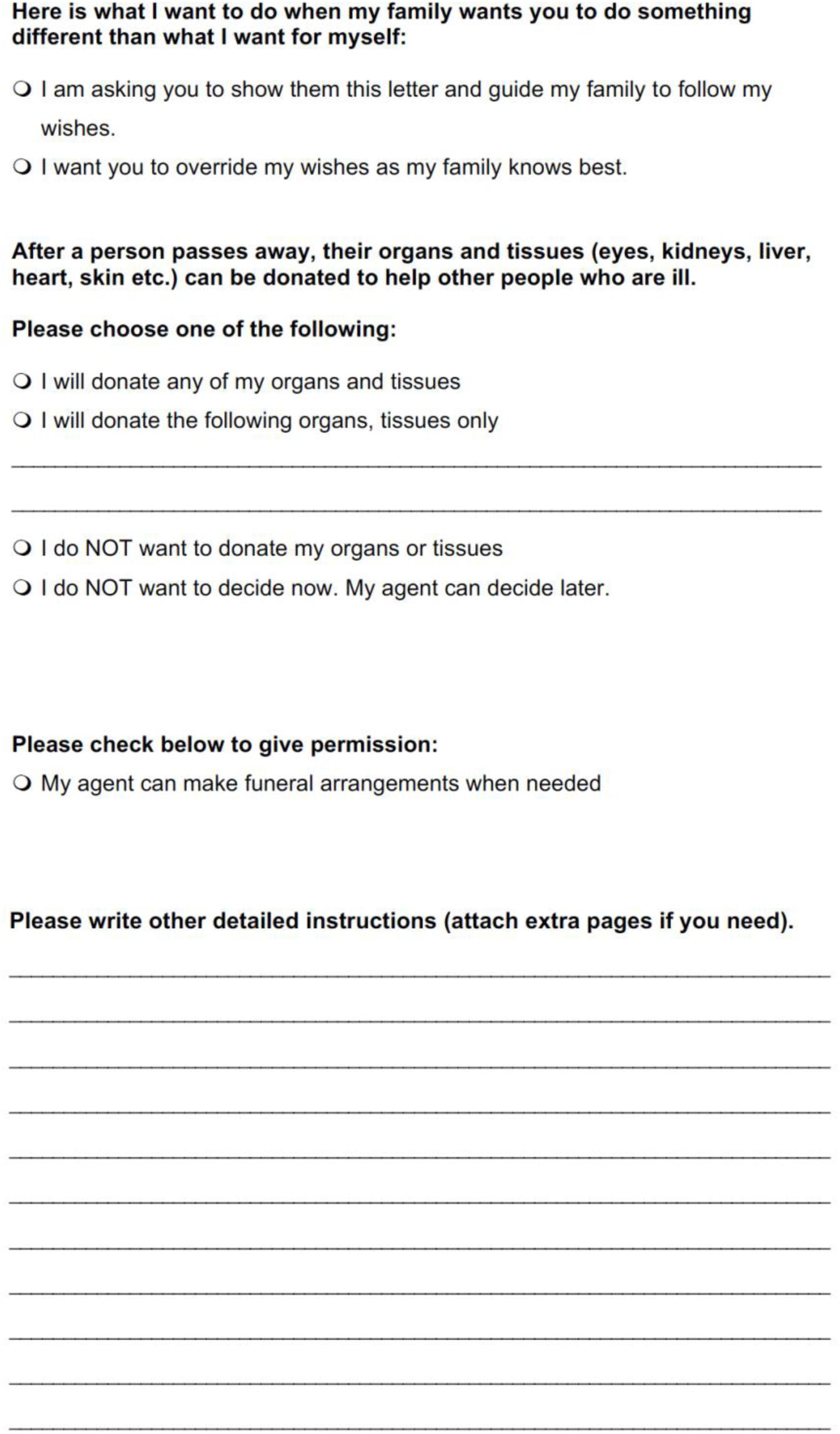

### Part 4: Sign the Form and have two witnesses co-sign

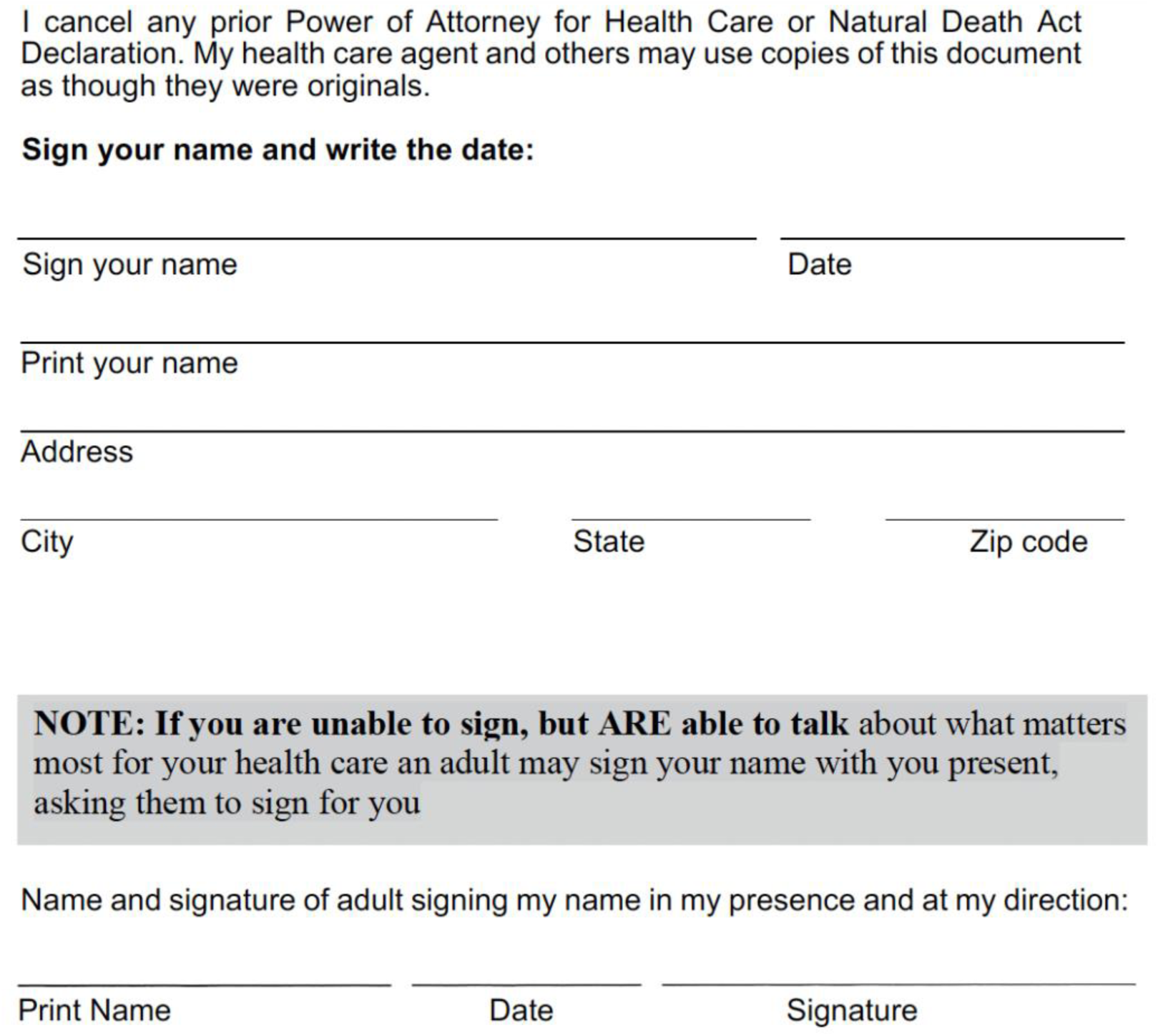
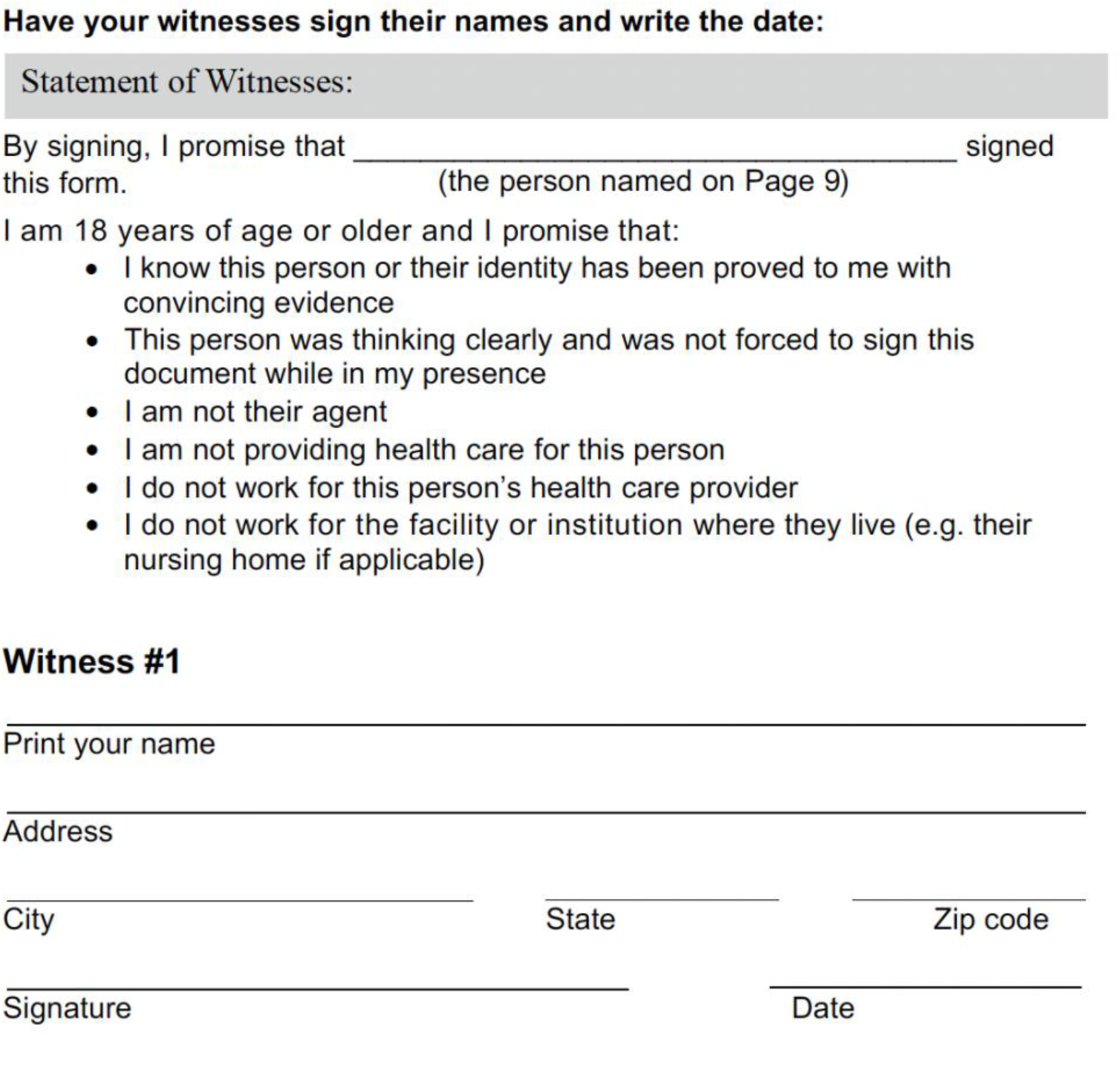
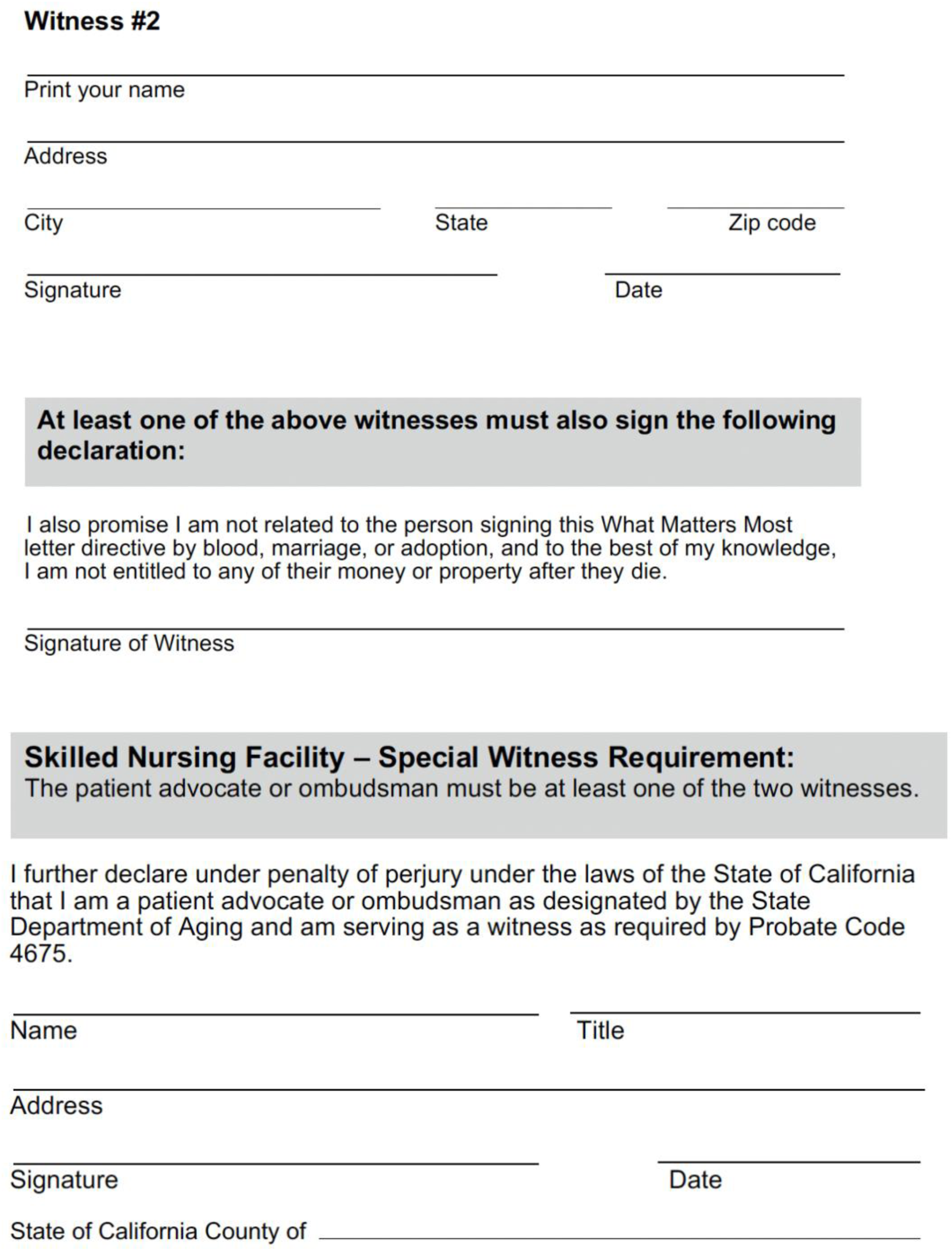

### Optional-Certificate of Acknowledgment of Notary Public

Signing before a notary public is not required if qualified witnesses have signed on pages 10-11. If you are a resident of a skilled nursing facility, you must have the patient advocate or ombudsman sign the statement of witnesses on pages 10-11 and the statement above, even if you choose to have this form notarized.

Signature__________________________(Seal)

On ________________ before me, ____________________ (insert name and title of officer), personally appeared __________________ (insert name of principal) who proved to me on the basis of satisfactory evidence to be the person(s) whose name(s) is/are subscribed to the within instrument and acknowledged to me that he/she/they executed the same in his/her/their authorized capacity(ies) and that by his/her/their signature(s) on the instrument the person(s), or the entity upon behalf of which the person(s) acted, executed the instrument.

I certify under PENALTY OF PERJURY under the laws of the State of California that the foregoing paragraph is true and correct.

WITNESS my hand and official seal.

### Part 5: Share a copy of this form with key people

- After you and witnesses have signed this form, please make photocopies and give to your health care agent, doctor, lawyer and other key people in your life.
- Ask your doctor to upload this form into your electronic medical record

## Resources

### Frequently Asked Questions

#### What is an advance health care directive?

An advance health care directive is a form that allows you to document how you want to be cared for in the future, if you cannot speak for yourself.

#### What is the Stanford What-Matters-Most letter directive?

The Stanford What-Matters-Most letter directive is a very simple type of advance health care directive that allows you to write a letter and tell your doctor and your family about what type of care you want in the future, if you cannot speak for yourself.

#### Who should write the What-Matters-Most letter directive?

Anyone 18 years or older should write their What-Matter-Most letter. If you have any health problems, you should definitely write your What-Matter-Most letter and tell your doctors, health team and your family about your wishes and health choices. Your doctors and your loved ones will be greatly relieved to know your wishes

#### Why should I write the What-Matters-Most letter directive?

Most doctors forget to talk to their patients about what type of care they want in the future and at the end of their life. Thus, patients do not have any say in what happens to them. By writing the Stanford What-Matters-Most letter, you will be able to tell your doctors and your family about the types of care you want.

#### Why should I write the What-Matters-Most letter directive *now?*

Some people wait until it is too late and never get to tell their doctors and health team about what matters most to them. The best time to write the letter is when you are healthy and able to speak for yourself.

#### What happens if I don’t write the What-Matters-Most letter directive?

When your doctors and health team do not know what matters most to you, they could unknowingly give you treatments that you may not want or treatments that may be too burdensome to you. If your loved ones do not know what you want, they will have to bear the burden of making decisions on your behalf (research has shown that this is very stressful for family members).

#### How do I know what treatment options are available to me?

Your doctors will tell you about your illness, give you information about any available treatments and their side effects. Your doctors can also tell you which treatments might be best for you but they cannot choose for you. You have to make decisions about your treatments based on what is important to you. If your doctors know what matters most to you, they will be able to help you with making decisions.

#### What do you want me to do?

You could answer the questions in the what-matters-most letter directive. Discuss it with your family and sign the letter directive in front of witnesses and give it to your doctor.

#### What does the word agent or proxy mean? Why do I need an agent (proxy)?

A healthcare agent, also known as a health care proxy is someone who will make medical decisions for you when you are not able to make your own decisions.

#### Who can be my agent?

Most people name a trusted friend or family member to make decisions in some situations, in the event they lose the ability to make their own decisions. You should name two or three people you trust to serve as your agent. If you do not name someone, then your healthcare team will choose an agent for you based on the regulations in the state you live in. An agent may also be referred to as a decisionmaker or proxy decisionmaker.

#### Who is NOT eligible to serve as my healthcare agent?

The following persons are NOT eligible by law to be your agent:

- Your doctors, nurses, and other health professionals taking care of you
- Any employees of the health facility where you are receiving care
- Any employees of a community care and residential care facility where you are receiving care.

#### When does my agent begin making my medical decisions?

In the event you are not able to make decisions for yourself anymore, your agent will make decisions on your behalf. However, if you wish, you can ask your agent to begin making decisions for you immediately

#### Should I tell my agent about my What-Matters-Most letter directive?

It is very important that you discuss your What-Matters-Most letter with your agent and help them understand your values and wishes. This way, when you are not able to make decisions for yourself, your agent can make decisions for you guided by your values and beliefs. The What-Matters-Most letter helps you describe your values to your agent. You should tell your agent that you have named them in your What-Matters-Most letter directive as your agent.

#### What if I don’t want to name a healthcare agent?

You do not have to name an agent, though it is in your best interest to do so. You can still write out your wishes and ask your doctor to list those wishes in your medical record. This will give your doctors and your family or friends some idea of what you would want if the time came that you couldn’t speak for yourself.

#### Will I still be treated if I don’t complete my What-Matters-Most letter directive?

Yes of course! We will always give you the best possible care. If you become too sick to make decisions yourself, someone else will have to make them for you.

#### What should I do after writing my What-Matters-Most letter directive?

Once you write and sign the letter, two qualified adults must witness your signature. Your doctors or members of your health team cannot witness your letter directive. If you prefer, you can notarize your letter directive.

#### What evidence of identity do you need to show the people who witness this form?

You can use any photo identification card issued by the United States federal government or state government (driver’s license, passport etc.) that is current or issued within the last five years. Other acceptable forms of ID include a current driver's license issued by the Canadian or Mexican government, or a valid foreign passport that has been stamped by the U.S. Immigration and Naturalization Service. Hospital issued identification (e.g. wrist band) can be used too.

#### What should I do after I have my What-Matters-Most letter directive signed and witnessed?

If you prepare the document at home, give copies of the form to your health care agent and anyone else of your family or friends who might be involved in your care. Please remember to bring a copy to your doctor at your next visit so it can be included in your hospital medical record. Make some extra copies so you can take one with you if you are admitted to a hospital, nursing home or any other health care facility. Keep the original in a place where you can access it easily and you can tell others how to find it as well. Once you have signed this What Matters Most Advance Directive form it will supersede or replace any earlier Advance Directive forms or Durable Power of Attorney for Health Care forms you have signed in the past.

#### Can I make changes to my What-Matters-Most letter directive?

You can change your What-Matters-Most letter directive at any time. You can also change your agent at any time. If you want to make changes, you must complete a new What-Matters-Most letter Directive, sign it and give it to your doctor to file in your hospital medical record.

#### In what languages is the What-Matters-Most letter directive available?

The What-Matters-Most letter is available in English and many languages like Spanish, Chinese, Tagalog, Vietnamese, Hindi, Urdu and others.

#### Can I make changes to my What-Matters-Most letter directive?

You can change your What-Matters-Most letter directive at any time. You can also change your agent at any time. If you want to make changes, you must complete a new What-Matters-Most letter Directive, sign it and give it to your doctor to file in your hospital medical record.

## Glossary of Terms

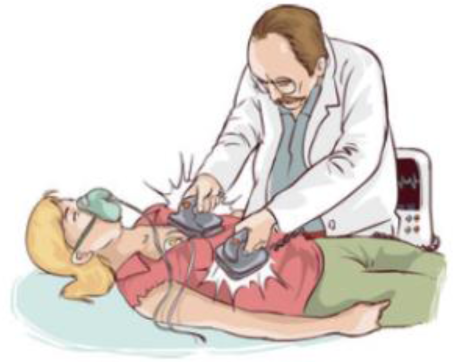

**Cardiopulmonary resuscitation (CPR)** – If a person’s heart stops or if that person stops breathing and the person has not indicated he or she does not want CPR, health care professionals usually try to revive him or her using CPR. In most cases when people have a terminal illness this is not successful. (You do not need to have an advance directive to request a do-not-resuscitate order.)

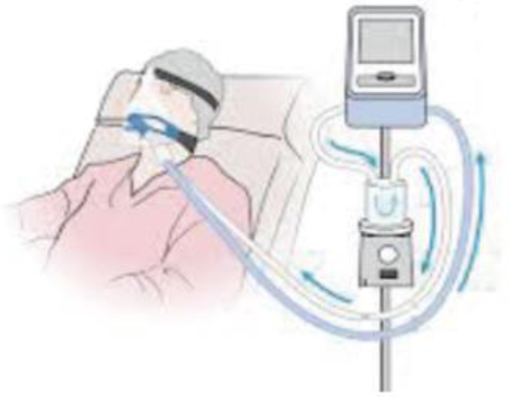

**Breathing machine** – If your lungs stop working properly, doctors can connect you to a machine called a ventilator. A ventilator is a machine that pumps air into a person’s lungs through a tube in the person’s mouth or nose that goes down the throat. The machine breathes for a person when he or she cannot.

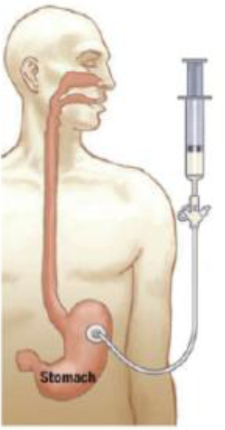

**Artificial liquid feeding** – There are various methods to feed people who can no longer eat, including inserting a tube into the stomach through a person's nose or through the stomach wall to give food and fluids.

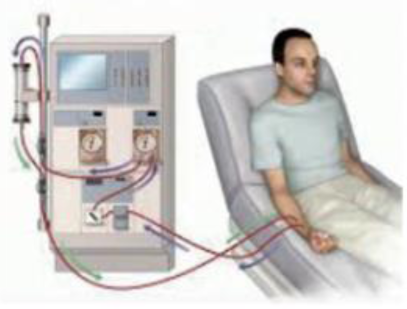

**Dialysis** – If your kidneys stop working properly, your blood can be cleaned using a dialysis machine, The dialysis machine does the work of your kidneys. Most people have to go to a dialysis center and be dialyzed three times a week. Some are dialyzed at home.

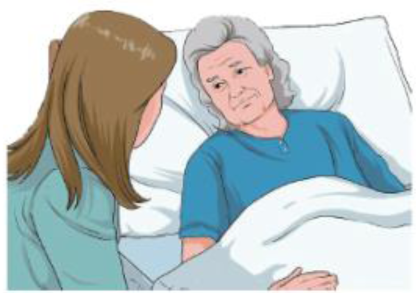

**Hospice care** – is a type of care provided to a patient at the end of life. Hospice care focuses on enhancing the dying person’s quality of life and provides support to their family or friends. Hospice care is usually provided in the home, but also can be provided in a hospital or nursing home.

## Who Will Help Me Complete this form

If you prepare the document while at Stanford Hospital, the staff who assist you will make copies and send a copy to Medical Records. You may also register your What Matters Most Directive with the California registry. Please refer to the following site for information: https://www.sos.ca.qov/reqistries/advance-health-care-directive-reqistrv/

### How can I get more information?

Please ask your doctor, nurse, social worker, chaplain or other member of the health team to help you. Additional copies of the What-Matters-Most letter directive are available from Spiritual Care Service (3-5101 from phones in the hospital or 650-723-5101 from phones outside the hospital)

### Are there other ways that I can make my health care wishes known?

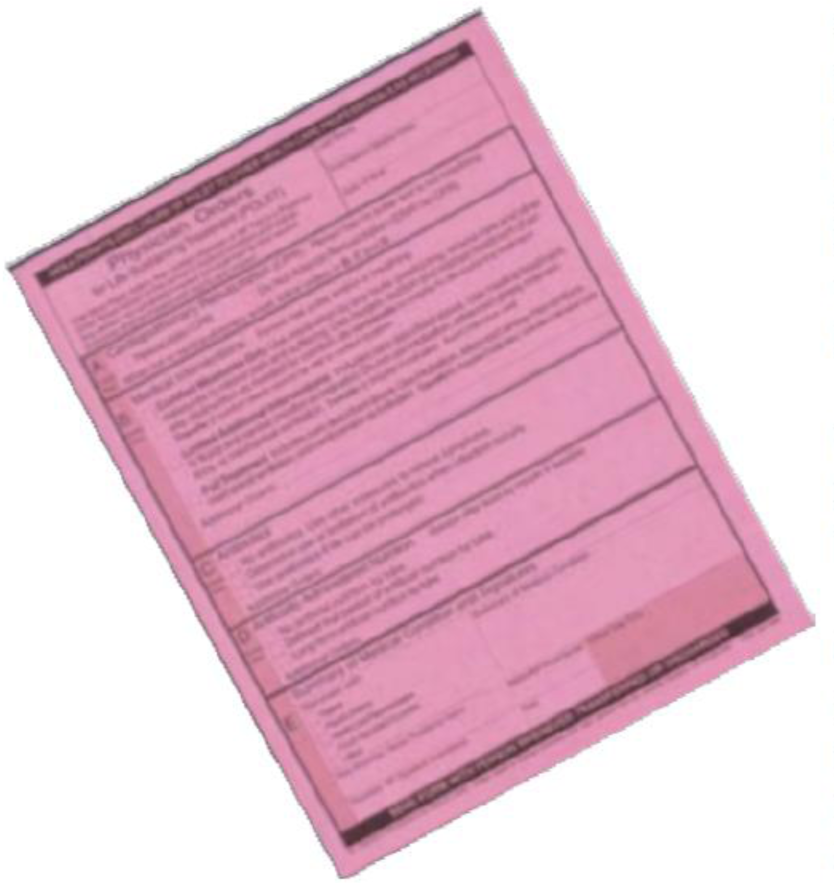

In addition to the What-Matters-Most letter directive, persons who are seriously ill can complete a POLST (Physician Orders for Life-Sustaining Treatment) form to express treatment preferences. It includes orders describing CPR, Medical interventions (intensity of care-ICU, no-ICU, comfort care, etc.) and the use of artificial nutrition. The POLST must be signed both by you and your doctor. It is not an Advance Directive and will not replace this What-Matters-Most letter directive. Information about the POLST is available at: http://www.capolst.org where you can also download a copy of the form.

### What if I already have an Advance Health Care Directive from another hospital? Can I use it?

Show the document to a social worker or chaplain to be reviewed and if it meets the basic criteria of an advance health care directive, it will be scanned into your medical record and used as indicated.

## Stanford Contact Information

If you are in the hospital and need help completing this form, contact Spiritual Care Service 650-723-5101, pager 15683. We take all feedback very seriously. Please direct your comments to Guest Services at 650-498-3333. For quality of care concerns, you may also contact the Joint Commission at 800-994-6610 or send an email to.

If you have any feedback specifically about the hospital’s provision of information on Advance Health Care Directives, you may contact:

Department of Health Services

Licensing and Certification Division

100 Paseo de San Antonio, Suite 235

San Jose, CA 95113

408-277-1784

FAX 408-277-1032

If you are a Medicare patient with any feedback about the hospital’s provision of information on Advance Health Care Directives, you may also call the Medicare hotline: 1-800-MEDICARE or 1-800-633-4227.

### PHONE NUMBER

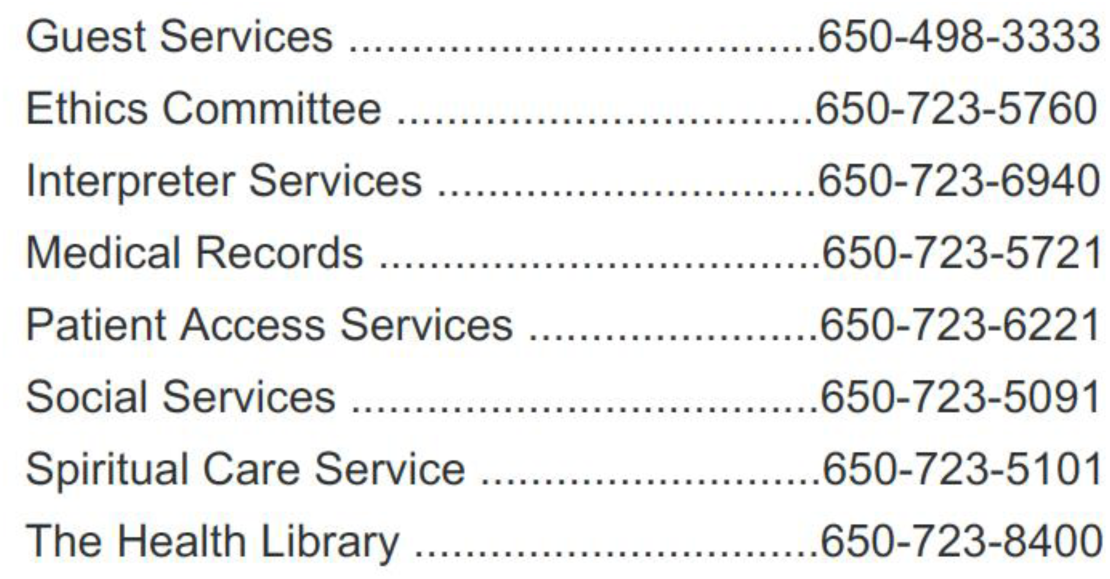

PHONE NUMBERS
|  |  |
| --- | --- |
| Guest Services ..... | 650-498-3333 |
| Ethics Committee ..... | 650-723-5760 |
| Interpreter Services ..... | 650-723-6940 |
| Medical Records ..... | 650-723-5721 |
| Patient Access Services ..... | 650-723-6221 |
| Social Services ..... | 650-723-5091 |
| Spiritual Care Service ..... | 650-723-5101 |
| The Health Library ..... | 650-723-8400 |

## Data Availability

Data are not available as the study is ongoing.

## Notes

### Competing Interest Statement

The authors have declared no competing interest.

### Clinical Trial

NCT03881579

### Funding Statement

NIA/NIH

### Author Declarations

This protocol was reviewed and approved by the Stanford University Institutional Review Board.

